# Microbiome signatures for detection of colorectal lesions in population-based FIT screening

**DOI:** 10.1101/2025.10.06.25336873

**Authors:** Einar E. Birkeland, Ane S. Kværner, Ekaterina Avershina, Cecilie Bucher-Johannessen, Vahid Bemanian, Hege S. Blix, Anette Hjartåker, Willem M. de Vos, Giske Ursin, Geir Hoff, Kristin R. Randel, Edoardo Botteri, Paula Berstad, Trine B. Rounge

## Abstract

The gut microbiome has been linked to colorectal cancer (CRC) development, with microbe-based classifiers distinguishing between CRC patients and healthy controls. However, there is a lack of studies addressing the utility of the microbiome in screening-relevant settings, including both precancers and CRC. In this Norwegian population-based study, we used fecal immunochemical test (FIT) leftovers from 1034 FIT-positive (i.e. positive for occult blood) screening participants for gut metagenome profiling using shotgun sequencing. Using comprehensive clinical, demographic, and lifestyle data, we modeled gut microbiome associations with CRC screening outcomes. Combining microbial profiles with quantitative FIT values improved detection of premalignant lesions beyond optimizing the FIT value alone, even after incorporating established CRC risk factors. Still, the FIT value maintained superior discriminative ability for CRC. We confirmed enrichment of bacteria such as *Fusobacterium nucleatum* and *Peptostreptococcus stomatis* in CRC. In contrast, other bacteria previously associated with the presence of CRC, including *Hungatella hathewayi* and *Clostridium symbiosum*, as well as *pks*-negative *Escherichia coli,* were enriched in those with no neoplastic findings, suggesting that their presence may reflect other conditions causing colon bleeding rather than underlying neoplasia. Microbial profiles were predominantly associated with distal rather than proximal lesions. Together, our findings highlight the potential for microbial markers to improve FIT-based CRC screening, especially by differentiating those with premalignant lesions from those who test FIT positive for other reasons.

## Introduction

The fecal immunochemical test (FIT), detecting fecal occult blood by measuring the concentration of human hemoglobin, has been widely adopted as the primary screening modality for organized colorectal cancer (CRC) screening programs^1^. In these programs, FITs are used to identify those who are more likely to have CRC or its precursors, thereby allocating constrained screening resources to those who will benefit the most. A positive FIT result is followed up with colonoscopy for detection of CRC and removal of its precursors. The FIT test is effective at identifying CRC, with a sensitivity of 0.75-0.91 for CRC^2^, and provides a reduction in CRC mortality compared to no systematic screening^3^. Still, the test has limited sensitivity for detecting pre-cancerous lesions^4^. Additionally, bleeding in the colon may also be caused by colonic conditions other than CRC or its precursors, such as hemorrhoids^5^ or inflammatory bowel disease^6^ (IBD) or other, unexplained reasons. Such conditions may cause FIT positivity, contributing to lower specificity for the FIT test in CRC screening.

To minimize the number of colonoscopy procedures carried out for screening participants who do not have CRC or precancers (false positives), the hemoglobin threshold used to determine colonoscopy referrals may be increased^7^. Increasing the threshold, however, entails a loss of sensitivity, especially for identification of individuals with precancerous neoplasia^8^. Therefore, the development of non-invasive biomarkers to complement or substitute bleeding-related markers may improve CRC screening.

The gut microbiome is a potential contributing factor in CRC development. Experimental evidence shows that transplantation of stool from CRC patients can promote carcinogenesis in mouse models^9^, and that use of antibiotics has been linked to increased risk of CRC^10^. Bacteria such as *pks*-positive *Escherichia coli* have been implicated in colorectal carcinogenesis^11^, whereas *Fusobacterium nucleatum* is often found in CRC tissues^12–14^. Studies of stool microbiomes from CRC patients and healthy controls have shown clear differences in the composition of microbes, indicating that the gut microbiome can be used for improved classification of CRC cases^15–18^.

The risk of developing CRC is affected by modifiable lifestyle factors. Key contributors include a sedentary lifestyle, adiposity, consumption of alcohol and red and processed meat, as well as smoking^19–22^. These lifestyle factors not only impact CRC risk, but several have also emerged as important modulators of microbiome diversity^23–25^. Additionally, non-modifiable factors such as age and sex impact both CRC risk and the gut microbiome^26^. Recent studies have shown how associations between the microbiome and CRC may be confounded by modifiable and non-modifiable factors^27,28^, emphasizing the importance of accounting for lifestyle factors when assessing the relationship between the microbiome and CRC.

So far, most microbiome studies have been conducted as case/control studies in the setting of established and symptomatic cancer contrasting healthy controls, with only a few studies including significant numbers of adenomas or other precancers^29,30^. Here, we evaluated the potential for microbiome-derived biomarkers in a Norwegian population-based CRC screening setting^31^, focusing on differentiating CRC or other colorectal neoplasms from FIT-positive individuals without neoplasia, a key challenge for complementing FIT in real-world screening. We analyzed gut microbiomes using shotgun metagenome sequencing of residual buffer from FIT-positive screening samples, integrating comprehensive data on lifestyle-related factors.

## Results

### Dataset description

To identify potential microbial biomarkers for the detection of participants at high risk for development of CRC, we performed metagenome sequencing using left-over buffer from FIT screening samples in the multicenter CRCbiome study in Norway^32^. The samples were originally collected as a part of a biennial population-based CRC screening pilot, the Bowel Cancer Screening Norway study^31^ (BCSN; cumulative participation rate 38.4% after three rounds of FIT). Those with a positive FIT test (>15 microgram Hb/g feces) were invited to participate in the CRCbiome study (N = 2700), and 1640 consented. Participants with findings of uncertain clinical significance (1-2 non-advanced adenomas, n = 406, or non-advanced serrated lesions, n = 104), who did not attend follow-up colonoscopy (n = 40) or for which sufficient sequencing data was not obtained (n = 49), were not included in the analyses (Supplementary Table 1). The study population thus consisted of 1034 individuals, where 582 were men (56%), and the median age at invitation was 67.1 years. In total, 584 (56.5%) had neoplastic findings (Table 1), including 66 (6.4%) individuals with CRC, 298 (28.8%) with advanced adenoma, 78 (7.5%) with advanced serrated lesions, and 142 (13.7%) individuals with three or more non-advanced adenomas as their most advanced finding (Supplementary Table S2). Participants without neoplastic findings (n = 450; 43.5%) included 9 individuals with “other” findings (inflammatory polyp, granulated tissue polyp), 272 with “non-neoplastic” findings (e.g. diverticulitis, hemorrhoids, IBD), and 169 with no findings.

**Table 1:**
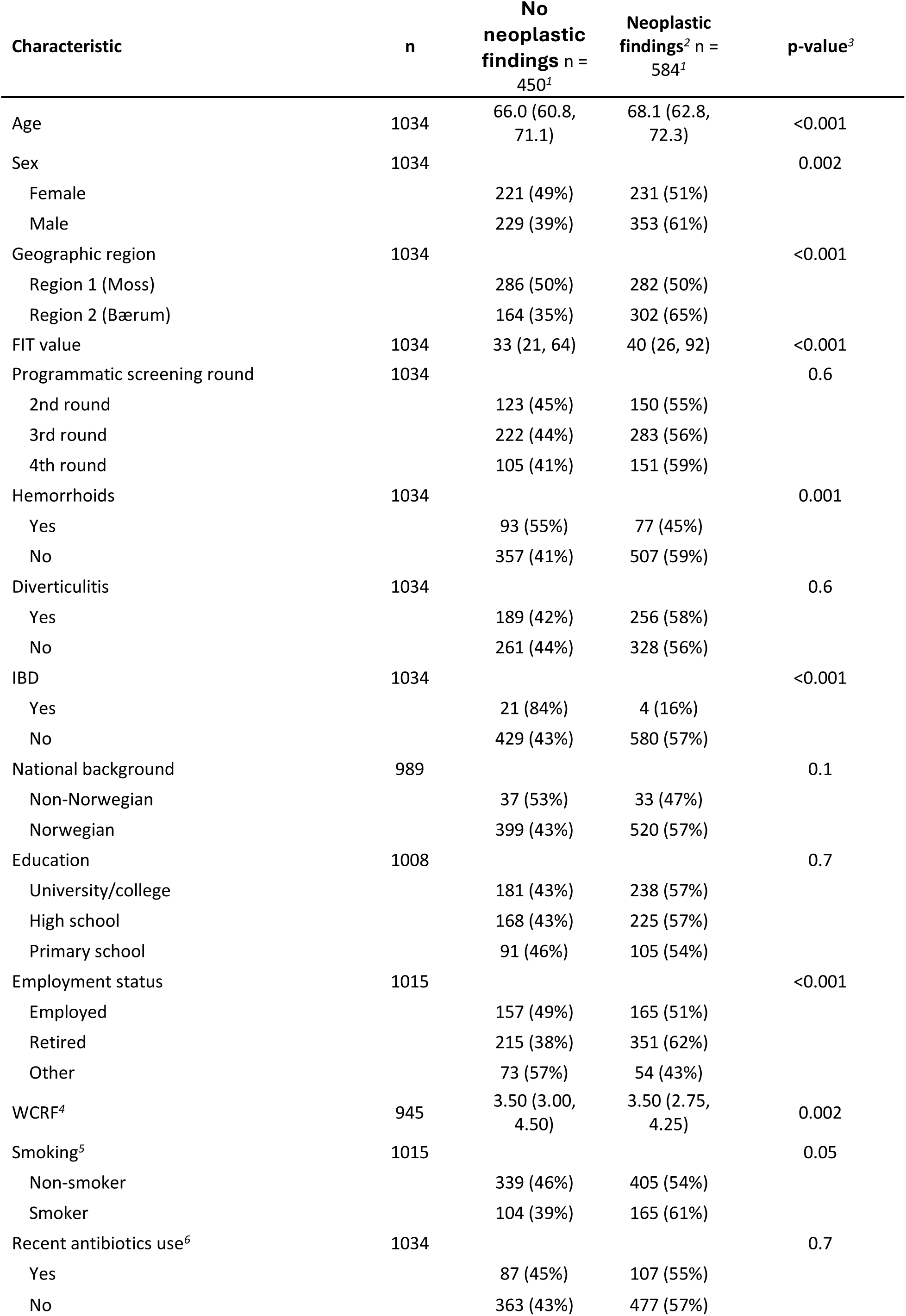
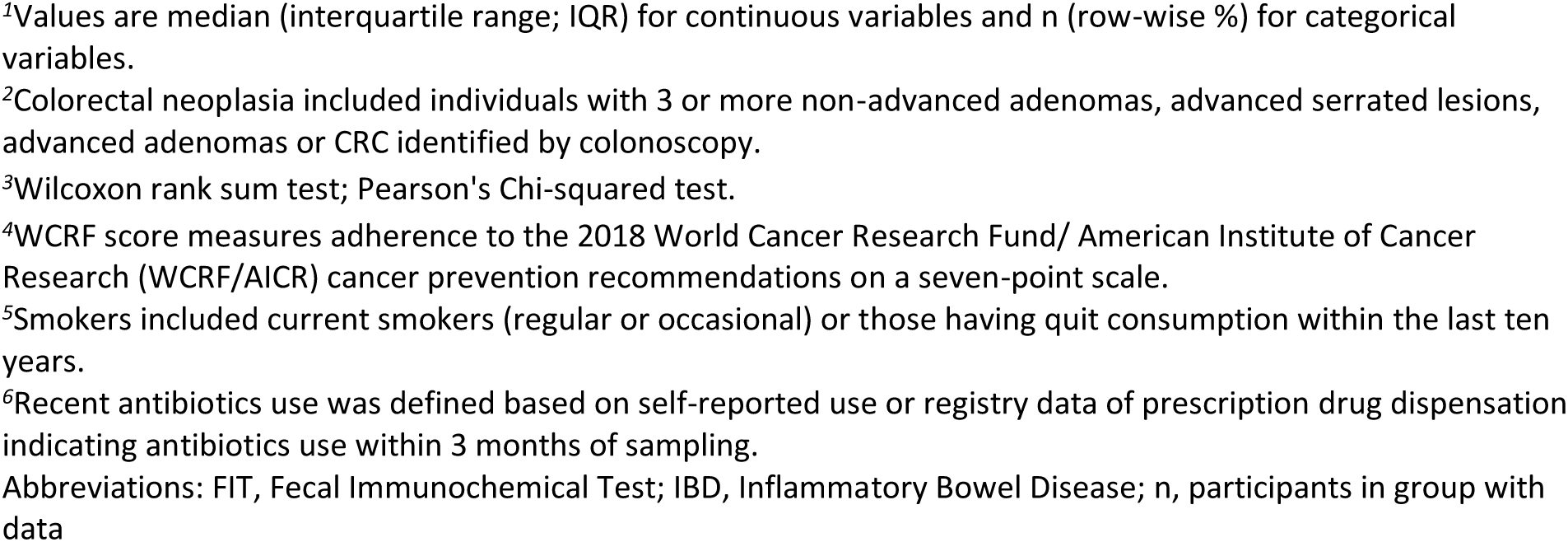
Participant characteristics.

| Characteristic | n | No<br>neoplastic<br>findings<sup>2</sup> n =<br>450<sup>1</sup> | Neoplastic<br>findings<sup>2</sup> n =<br>584<sup>1</sup> | p-value<sup>3</sup> |
| --- | --- | --- | --- | --- |
| Age | 1034 | 66.0 (60.8,<br>71.1) | 68.1 (62.8,<br>72.3) | <0.001 |
| Sex | 1034 |  |  | 0.002 |
| Female |  | 221 (49%) | 231 (51%) |  |
| Male |  | 229 (39%) | 353 (61%) |  |
| Geographic region | 1034 |  |  | <0.001 |
| Region 1 (Moss) |  | 286 (50%) | 282 (50%) |  |
| Region 2 (Bærum) |  | 164 (35%) | 302 (65%) |  |
| FIT value | 1034 | 33 (21, 64) | 40 (26, 92) | <0.001 |
| Programmatic screening round | 1034 |  |  | 0.6 |
| 2nd round |  | 123 (45%) | 150 (55%) |  |
| 3rd round |  | 222 (44%) | 283 (56%) |  |
| 4th round |  | 105 (41%) | 151 (59%) |  |
| Hemorrhoids | 1034 |  |  | 0.001 |
| Yes |  | 93 (55%) | 77 (45%) |  |
| No |  | 357 (41%) | 507 (59%) |  |
| Diverticulitis | 1034 |  |  | 0.6 |
| Yes |  | 189 (42%) | 256 (58%) |  |
| No |  | 261 (44%) | 328 (56%) |  |
| IBD | 1034 |  |  | <0.001 |
| Yes |  | 21 (84%) | 4 (16%) |  |
| No |  | 429 (43%) | 580 (57%) |  |
| National background | 989 |  |  | 0.1 |
| Non-Norwegian |  | 37 (53%) | 33 (47%) |  |
| Norwegian |  | 399 (43%) | 520 (57%) |  |
| Education | 1008 |  |  | 0.7 |
| University/college |  | 181 (43%) | 238 (57%) |  |
| High school |  | 168 (43%) | 225 (57%) |  |
| Primary school |  | 91 (46%) | 105 (54%) |  |
| Employment status | 1015 |  |  | <0.001 |
| Employed |  | 157 (49%) | 165 (51%) |  |
| Retired |  | 215 (38%) | 351 (62%) |  |
| Other |  | 73 (57%) | 54 (43%) |  |
| WCRF<sup>4</sup> | 945 | 3.50 (3.00,<br>4.50) | 3.50 (2.75,<br>4.25) | 0.002 |
| Smoking<sup>5</sup> | 1015 |  |  | 0.05 |
| Non-smoker |  | 339 (46%) | 405 (54%) |  |
| Smoker |  | 104 (39%) | 165 (61%) |  |
| Recent antibiotics use<sup>6</sup> | 1034 |  |  | 0.7 |
| Yes |  | 87 (45%) | 107 (55%) |  |
| No |  | 363 (43%) | 477 (57%) |  |

<sup>1</sup>Values are median (interquartile range; IQR) for continuous variables and n (row-wise %) for categorical variables.
<sup>2</sup>Colorectal neoplasia included individuals with 3 or more non-advanced adenomas, advanced serrated lesions, advanced adenomas or CRC identified by colonoscopy.
<sup>3</sup>Wilcoxon rank sum test; Pearson's Chi-squared test.
<sup>4</sup>WCRF score measures adherence to the 2018 World Cancer Research Fund/ American Institute of Cancer Research (WCRF/AICR) cancer prevention recommendations on a seven-point scale.
<sup>5</sup>Smokers included current smokers (regular or occasional) or those having quit consumption within the last ten years.
<sup>6</sup>Recent antibiotics use was defined based on self-reported use or registry data of prescription drug dispensation indicating antibiotics use within 3 months of sampling.
Abbreviations: FIT, Fecal Immunochemical Test; IBD, Inflammatory Bowel Disease; n, participants in group with data

Individuals with neoplastic findings had higher concentrations of blood in their stools (FIT-value) than those without (Table 1, Supplementary Table S2) and were less likely to have hemorrhoids and IBD. In line with the current consensus on risk factors for CRC^33^, neoplastic findings were more frequently detected in men, participants of advanced age, smokers, and those with lower adherence to cancer prevention recommendations (the WCRF score; see methods). Previously we have also shown that participants with colorectal neoplasia have higher intake of alcohol and red and processed meat^20,22^. Neoplastic findings were more frequent in individuals from region 2 (Bærum; 302/466 (65%)) than from region 1 (Moss; 282/568 (50%)).

To evaluate the representativeness of the CRCbiome study population, we compared their use of prescription drugs for common comorbidities with that of the full FIT arm of the BCSN study (N = 46 962), and a nationally representative screening-eligible population (N = 16 500) (see Methods). Drug use for the CRCbiome population within 1 year of initial invitation to BCSN indicated a higher rate of cardiovascular disease (53% vs 46%) and more frequent use of antibiotics (28% vs 26%) for CRCbiome participants, than the general screening eligible population (Supplementary Table S3). Notably, comorbidity-related prescription drug use did not differ substantially from the BCSN FIT arm.

### Microbial diversity and composition in a CRC screening population

Read-based taxonomic and functional profiling resulted in identification of 2756 microbial species, with a mean of 239 identified per sample (range = 68–498), and a mean of 292 162 (SD = 59 300) gene families per sample, aggregated to a mean of 2678 (SD = 564) KEGG orthogroups (KOs). Gut microbial profiles were dominated by Bacillota (former Firmicutes) species, both in prevalence, relative abundance, and species diversity (Figure 1a-c; Supplementary Figure 1a-b). The FIT-derived taxonomic profiles were consistent with publicly available gut microbiomes from stool samples^34^ (Supplementary Figure S2), and an assembly-based approach resulted in similar microbial profiles (Supplementary Figure S3). All results are thus reported using read-based profiles only. Microbial diversity (represented by number of observed species and KO groups, the Shannon and inverse Simpson indices), was significantly associated with factors related to CRC risk and socioeconomic status, including physical activity, adherence to cancer prevention recommendations (WCRF score), alcohol consumption, age, smoking status, recent use of antibiotics (within 3 months; self-reported or as indicated by prescription drug records) (Figure 1d, Supplementary Data 1), education level, and employment status. Similar observations were made for microbial composition where WCRF score showed the greatest explanatory power, with a PERMANOVA R² of 0.0067 (p < 0.001), followed by smoking status, body mass index (BMI), and education level (Figure 1e, Supplementary Data 2).

**Figure 1:**
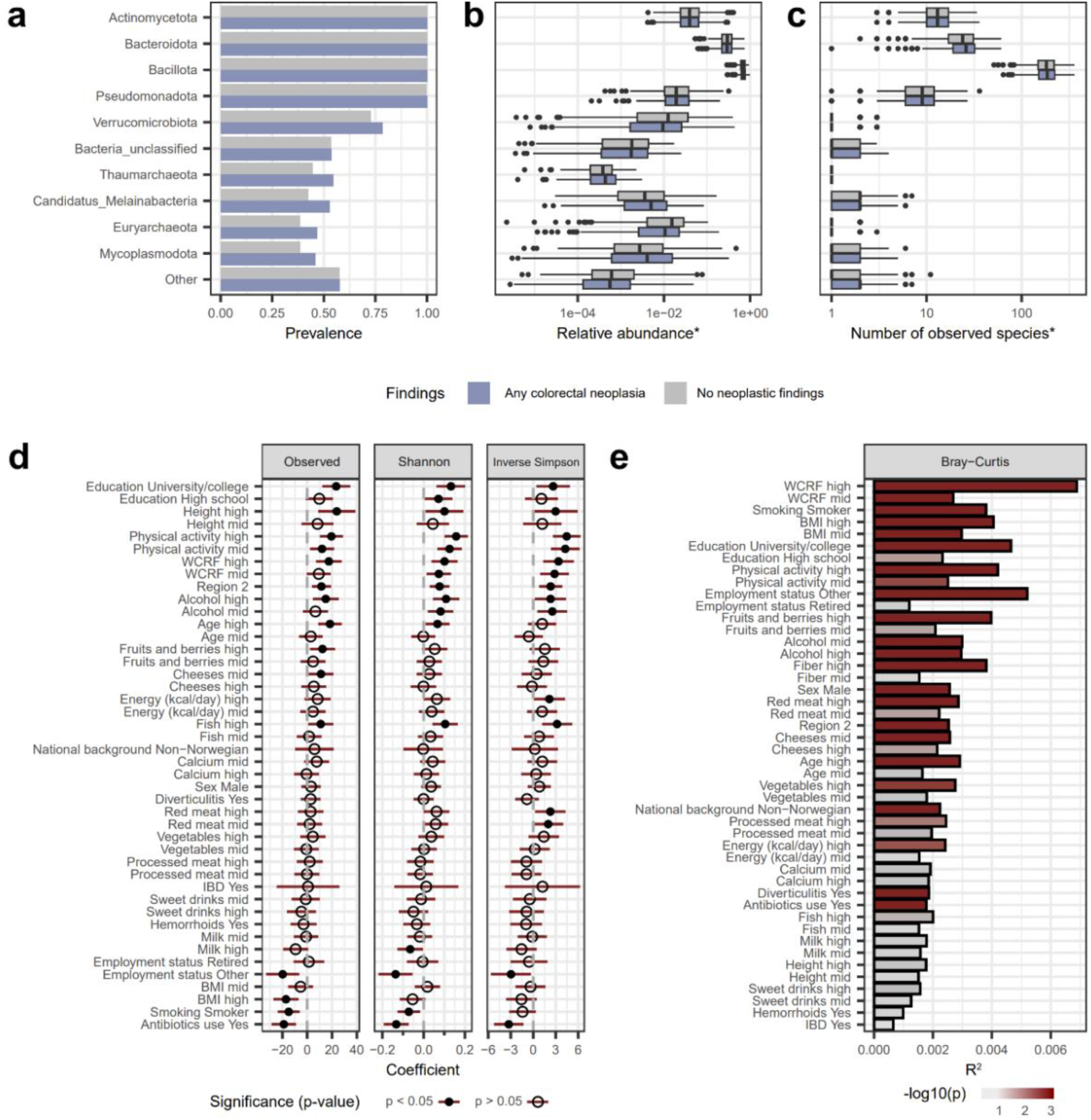
Microbial diversity in fecal immunochemical tests from screening participants. a) Prevalence and b) relative abundance of the 10 most prevalent microbial phyla (with remaining phyla grouped in “other”) stratified by participant colonoscopy findings defined as CRC-related (blue) or not (gray). For visualization purposes, relative abundance of zero has been omitted. c) The number of species observed per microbial phylum. d) Coefficients for associations between microbial alpha diversity measured as observed species, Shannon diversity, and inverse Simpson diversity and participant lifestyle and demographic factors. Associations were evaluated using a linear model, adjusting for participant age, sex, geographic region, and sequencing reads. Continuous variables were split into tertiles, with the lowest tertile considered the reference; coefficients denote the difference in diversity associated with variable level relative to reference. Participants with unknown or missing data were excluded, as were categories of participants with 10 or fewer observations. Lines indicate 95% confidence intervals. Significant differences are represented by black dots. e) Associations between gut microbiome beta diversity (Bray-Curtis distances based on relative species abundance) and lifestyle and demographic factors. Bars show variance explained by participant characteristics in permutational analysis of variance (PERMANOVA) models, in pairwise comparisons between levels of each variable: continuous variables were split in tertiles, and tests were made comparing each level to the lower tertile. Participants with unknown or missing data were excluded, as were categories of participants with 10 or fewer observations. Bars are shaded by PERMANOVA significance from light gray (p = 1) to dark red (p = 0.001). * Abundance or diversity values of 0 were omitted from b) and c).

Microbial associations with clinicopathological outcomes were evaluated in models adjusted for lifestyle, diet, antibiotics use, and gastrointestinal conditions (see methods). Here, each standard deviation increase in Shannon diversity was associated with 16% higher odds of detecting any colorectal neoplasia (OR = 1.16 [1.01-1.33]; Figure 2a), and specifically 24% for detection of advanced adenomas relative to no neoplastic findings (OR = 1.24 [1.05-1.47]). Conversely, colorectal neoplasia detection was negatively associated with the number of KO-groups (OR = 0.78 [0.68-0.90]). No significant associations were found for those with CRC or advanced serrated lesions.

**Figure 2:**
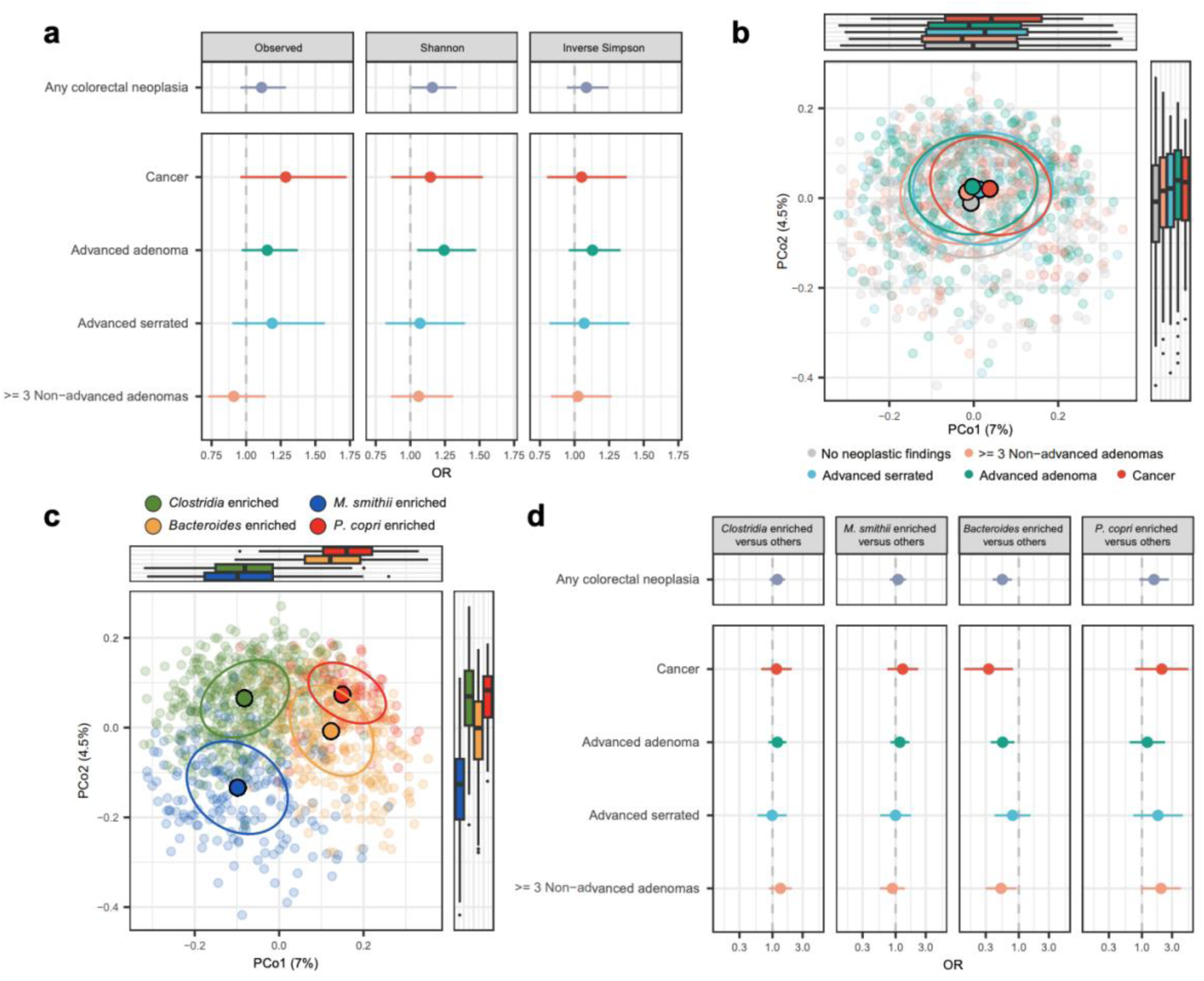
Associations between outcome groups and microbial diversity, composition, and community states. a) Odds ratios of outcome groups associated with one standard deviation increase in microbial alpha diversity, estimated using multinomial logistic regression models adjusted for sequencing depth, age, sex, geographic region, smoking, level of education, the WCRF score, hemorrhoids, IBD, and recent use of antibiotics. b) PCoA based on Bray Curtis distances, categorized by diagnostic groups. Solid points indicate centroids, and ellipses encompass 50% of observations. c) PCoA as in b) categorized by microbial community states derived from Dirichlet multinomial mixture models. d) Odds ratios of outcome groups associated with having one microbial community state relative to any other, presented in panels from left to right. Models were adjusted as in a), except for not including sequencing depth.

Compositional differences were found between those with and without neoplastic findings (PERMANOVA R^2^ = 0.002, p = 0.002), and between diagnostic groups (PERMANOVA R^2^ = 0.005, p = 0.011; Figure 2b). The largest differences were observed between participants with no neoplastic findings and those with advanced adenomas (Supplementary Figure S4; Pseudo F = 1.60). Using Dirichlet multinomial mixture models (DMMs^35^; Figure 2c) microbial profiles were classified into four community states with the largest groups enriched by *Clostridium* spp. (n = 425); *Methanobrevibacter smithii* (n = 331); *Bacteroides* spp. (n = 200); or *Prevotella copri* (n = 78; Supplementary Figure S5a). Gut community states were associated with several host features, including sex, the WCRF score, and recent anti-acid medication use (Supplementary Figure S5b).

Notably, neoplastic findings were less common among those with a *Bacteroides*-enriched community state (OR = 0.55 [0.39-0.78]; Figure 2d, Supplementary Table 4), specifically, these participants were less likely to be diagnosed with CRC (OR = 0.33 [0.14-0.81]). An opposite non-significant trend was observed for the community state dominated by *P. copri* (OR = 2.0 [0.78-5.44]).

### Prediction of colorectal neoplasia using microbial models

To assess the utility of the microbiome as a potential biomarker for colonoscopy need, we constructed machine learning models for identification of individuals with neoplastic findings. Here, 20% of the dataset was set aside as a test set for validation purposes. The random forest (RF) machine learning algorithm with relative abundance of microbial species as predictors outperformed other classification methods and gene content (KOs) as predictors, whether used alone or in combination with demographic and lifestyle factors (see Supplementary Figure S6).

The best-performing microbe-based RF models predicted neoplastic findings with a mean AUROC across 20 times repeated 5-fold cross-validation of 0.610 (SD = 0.036) in the training set and 0.56 (95% CI = 0.48-0.64) in the test set (Table 2). The microbe-based model demonstrated improved predictive performance for identifying colorectal neoplasia relative to raising the FIT threshold above 15 µg Hb/g feces in the training set (FIT AUROC = 0.564, SD = 0.042), but more ambiguous results in the test set (FIT AUROC = 0.60, 95% CI = 0.52-0.68). We employed an ensemble approach for prediction of neoplastic findings, incorporating both microbial predictions and the FIT value, taking advantage of the FIT accuracy for CRC, achieving an AUROC of 0.615 (SD = 0.040) in the training set and 0.61 (95% CI = 0.54-0.69) in the test set. In ensemble models, the microbial predictions contributed to model accuracy (median LRT p-value < 0.001 [max p-value = 0.004]), and were generally more accurate in the 20% cross-validation sets for the combined FIT-microbial models than for the FIT alone. However, these differences in accuracy did not consistently reach statistical significance (median AUROC difference = 0.05, median p-value for AUROC difference = 0.25, significant differences in 20% of repeats).

**Table 2:** Training and test set prediction accuracy for neoplastic findings.

| Model | AUROC<sup>1</sup> | Sensitivity<sup>1</sup> | Specificity<sup>1</sup> |
| --- | --- | --- | --- |
| *Training set* |  |  |  |
| FIT alone | 0.564 (0.042) | 0.594 (0.133) | 0.496 (0.141) |
| Microbial RF model alone | 0.610 (0.036) | 0.810 (0.115) | 0.322 (0.153) |
| Microbial RF model + FIT | 0.615 (0.040) | 0.754 (0.126) | 0.398 (0.163) |
| Microbial RF model + FIT, sex, age, and WCRF score | 0.639 (0.037) | 0.835 (0.089) | 0.328 (0.117) |
| FIT, sex, age, and WCRF score | 0.607 (0.037) | 0.806 (0.122) | 0.332 (0.144) |
| *Test set* |  |  |  |
| FIT alone | 0.60 (0.52-0.68) | 0.81 (0.74-0.88) | 0.32 (0.22-0.42) |
| Microbial RF model alone | 0.56 (0.48-0.64) | 0.78 (0.70-0.84) | 0.31 (0.22-0.41) |
| Microbial RF model + FIT | 0.61 (0.54-0.69) | 0.82 (0.75-0.89) | 0.29 (0.20-0.38) |
| Microbial RF model + FIT, sex, age, and WCRF score | 0.64 (0.56-0.72) | 0.82 (0.74-0.89) | 0.38 (0.27-0.49) |
| FIT, sex, age, and WCRF score | 0.61 (0.53-0.69) | 0.77 (0.69-0.85) | 0.31 (0.21-0.41) |
| <sup>1</sup> Metric uncertainty given as standard deviation across 20 times repeated 5-fold cross-validation for the training set, and as 95% confidence intervals for 1000-fold bootstrap for the test set |  |  |  |

When including participant age, sex, and the WCRF score as predictors, the accuracy for prediction of neoplastic findings improved, with a mean AUROC in the training set of 0.639 (SD = 0.037), and 0.64 (95% CI = 0.56-0.72) in the test set. Notably, omitting the microbial score from the predictors resulted in a decline in prediction accuracy (training AUROC = 0.607; SD = 0.037, test AUROC = 0.61, 95% CI = 0.53-0.69). Microbial predictions contributed to ensemble model accuracy also in this case (median LRT p-value < 0.001). At a threshold where 90% of CRC cases were correctly classified, microbial models exhibited higher sensitivity for the detection of precancerous lesions than the FIT value alone, although use of the FIT value alone had higher specificity (Figure 3a, right panel; Table 2).

**Figure 3:**
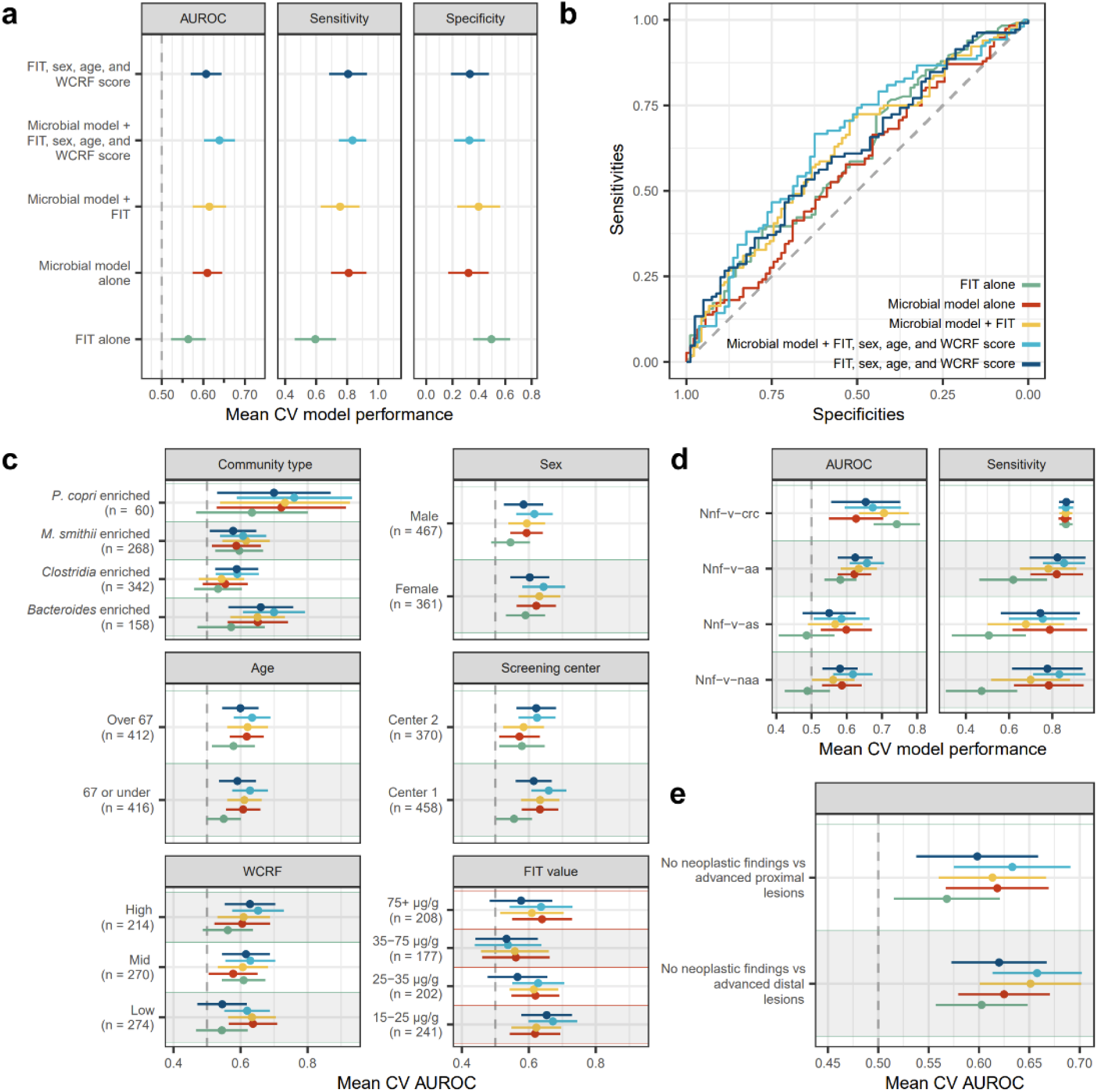
Prediction of colorectal neoplasia based on gut microbial species abundance. a-e) Random forest models based on microbial species abundance were constructed and combined with either the FIT value or the FIT value, sex, age, and WCRF score using logistic regression. Results are also presented for the FIT value alone as a predictor, or for a model including the FIT value, participant age, sex, and WCRF score, with colors indicating models as in a). a) Classification results from the training set are summarized as the mean (± SD) of 20-times repeated 5-fold cross-validation (CV) of AUROC, sensitivity, and specificity, in panels from left to right. Sensitivity and specificity were defined as the probability score resulting from setting the threshold for positivity to include 90% of CRC cases. b) Receiver operating characteristic curves for model predictions in the test set (20% of the dataset; n = 206). c) Model performance in the training set for participant subsets. The number of participants in the training set for each subset is indicated. d) Model performance for classification of colorectal neoplasia by colonoscopy-based diagnosis relative to no neoplastic findings. Sensitivity was measured using the same cut-off as in a), and specificity is omitted, as it is identical to that reported in a). e) Predictive performance for colorectal neoplasia stratified by lesion localization. Only advanced lesions were treated as cases, and the comparison was made only for those without a corresponding lesion in the opposite compartment (Those with advanced distal lesions were excluded from the comparison of proximal lesions to those with no neoplastic findings (nnf), and vice versa). Abbreviations: aa, advanced adenoma; as, advanced serrated; crc, colorectal cancer; naa, non-advanced adenoma; nnf, no neoplastic findings.

Classification accuracy using microbiome-based models was relatively stable across subsets of participant characteristics (Figure 3c and Supplementary Figure S7a). Overall, there was a slight improvement in prediction accuracy among participants without recent use of antibiotics, who belonged to region 1 (Moss), were female, and in the lower ranges of FIT values. The largest impact on prediction accuracy was associated with microbial community state, where among those with enrichment of *P. copri*, the microbial model had an AUROC of 0.72 (SD = 0.19). Combining the microbiome-based predictor with either the FIT value or with the FIT value, sex, age, and WCRF score, consistently improved prediction accuracy for colorectal neoplasia across participant stratifications except for those with drug use indicating diabetes, or those who had recently used antibiotics (Figure 3c and Supplementary Figure S7a).

For classification of neoplastic findings, feature selection and subsampling of the training set showed increased predictive potential associated with inclusion of more features but only limited improvements with increased sample size (Supplementary Figure S7b).

We investigated whether microbiome-informed models could improve prediction of specific types of premalignant lesions (Figure 3d). For CRC, the FIT value had slightly higher predictive power (mean AUROC = 0.742) than the combined microbial/FIT model (mean AUROC = 0.715). Notably, both the microbial model alone, and in combination with the FIT value, were better able to detect precancerous neoplastic findings than the FIT alone at varying thresholds for positivity (Supplementary Figure S8). Evaluation of lesion detection by localization showed increased accuracy associated with addition of the microbial model for both distally and proximally localized lesions (Figure 3e).

### Differential abundance between outcome groups

To identify individual microbes associated with colorectal neoplasia, we carried out multivariate adjusted differential abundance analyses (see methods) including species present in at least 10% of participants. Here, 13 species were differentially abundant between those with and without colorectal neoplasia (Figure 4a, left panel; Supplementary Data 3), and 20 species were differentially abundant when contrasting any of the diagnostic groups to those without neoplastic findings (Figure 4a, right panel). In unadjusted models, only 9 species were associated with neoplastic findings, and 10 with any specific outcomes, indicating that statistical adjustment was necessary to uncover significant associations with outcome groups. For KOs, 25 and 11 were significantly associated with colorectal neoplasia or any diagnostic group, respectively (Supplementary Data 4).

**Figure 4:**
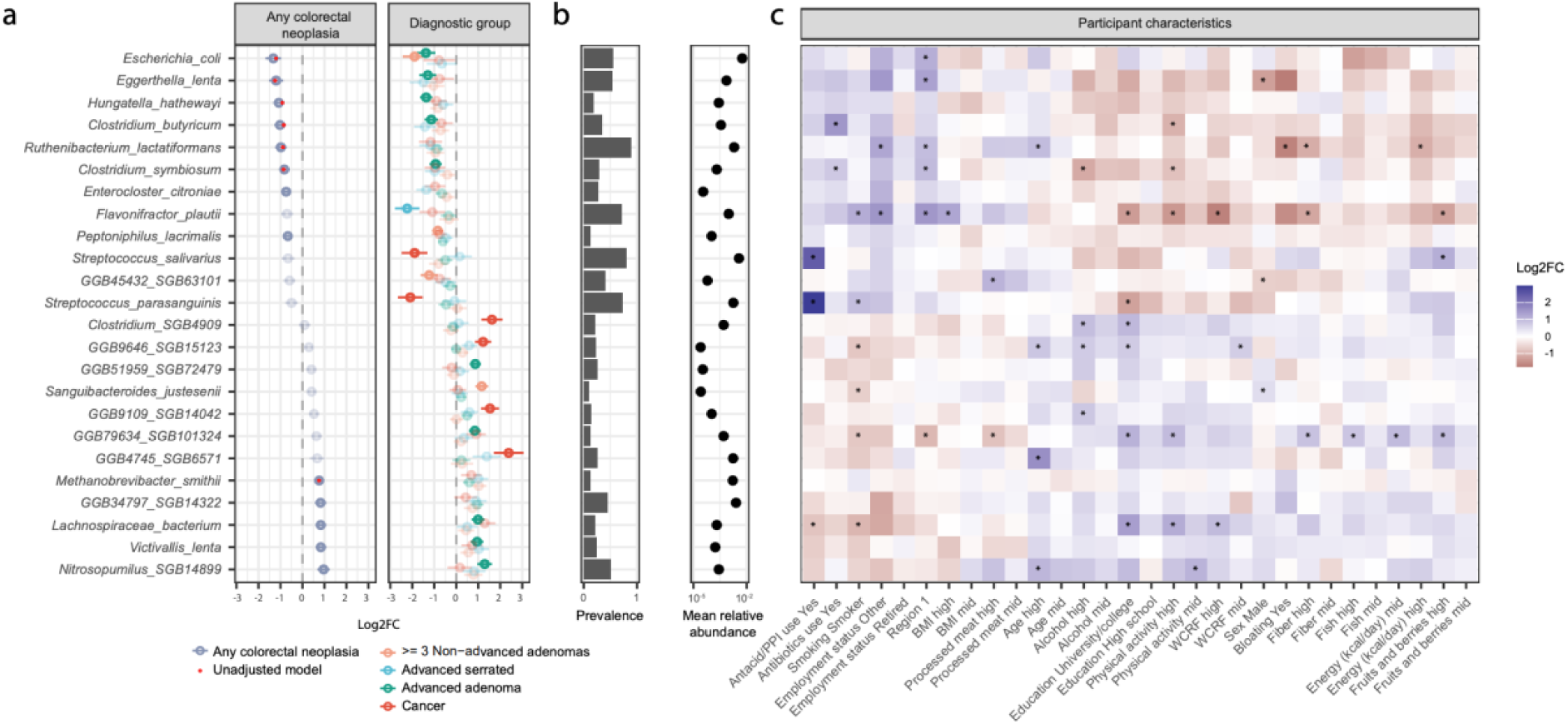
Differential abundance of species between diagnostic groups. **a)** Effect sizes of differential abundance of microbial species for those with relative to those with no neoplastic findings (left panel) or for each diagnostic group relative to those with no neoplastic findings (right panel). Only species differentially abundant with regards to colorectal neoplasia, or for CRC, advanced adenomas, or advanced serrated lesions relative to no neoplastic findings are included in the plot (a full list of differentially abundant species is available in Supplementary Data 3). Differential abundance models included adjustment for age, sex, geographic region, sequencing reads, smoking status, level of education, the WCRF score, hemorrhoids, IBD and recent use of antibiotics. Circles indicate the log2 fold change and lines standard error. Opaque symbols indicate significant associations at FDR < 0.05, and semi-transparent ones indicate an FDR value >0.05. For colorectal neoplasia, log2 fold changes derived from unadjusted models are shown as red points. **b)** Prevalence (left panel) and mean relative abundance (right panel) of differentially abundant species as in a). **c)** Species with significant associations with diagnostic groups were evaluated for associations with participant characteristics in separate models (adjusted for participant age, sex, and region). Colors show coefficient magnitude and direction, with significant associations indicated by asterisks. Participant characteristics measured on a continuous scale were split in tertiles (low, mid and high), where low was considered the reference value. Reference values for categorical variables were as follows: antacid/antibiotic use – No; smoking status – non-smoker; region – Region 2; and employment status – employed. Only variables with any significant associations are displayed; a full table of results is available in Supplementary Data 5.

Comparing species abundance between pairs of diagnostic groups indicated incremental changes in microbial communities from those with no neoplastic findings through stages of CRC precursors, to CRC, with the clearest contrast between those with no neoplastic findings and those with CRC (Supplementary Figure 9).

Species with differential abundance did not display any consistent pattern in overall prevalence or abundance (Figure 4b); however, they were frequently associated with host characteristics, (Figure 4c; Supplementary Data 5). In general, microbes positively associated with colorectal neoplasia were typically also positively associated with beneficial factors such as the level of physical activity.

*E. coli* abundance was associated with having no neoplastic findings (Figure 4a), and the presence of *E. coli* (n = 568; 55%) was associated with a 43% lower odds of colorectal neoplasia (OR = 0.57 [95% CI = 0.43-0.75]; Figure 5a). As previous research has shown *pks* positive *E. coli* to be associated with CRC^36^, we stratified samples with *E. coli* by the presence of the *pks* gene cluster, finding 167 samples to be *pks* positive (>10% coverage of the colibactin gene cluster by read mapping), and 401 to be *pks* negative. Interestingly, *pks*-negative *E. coli* was associated with a lower rate of colorectal neoplasia (OR = 0.53 [0.39-0.71]), and a contrasting trend towards a higher rate of colorectal neoplasia, and of CRC in particular, among those with pks-positive *E. coli* (OR = 1.33 [0.89-1.98] and OR = 1.99 [0.94-4.20], respectively).

**Figure 5:**
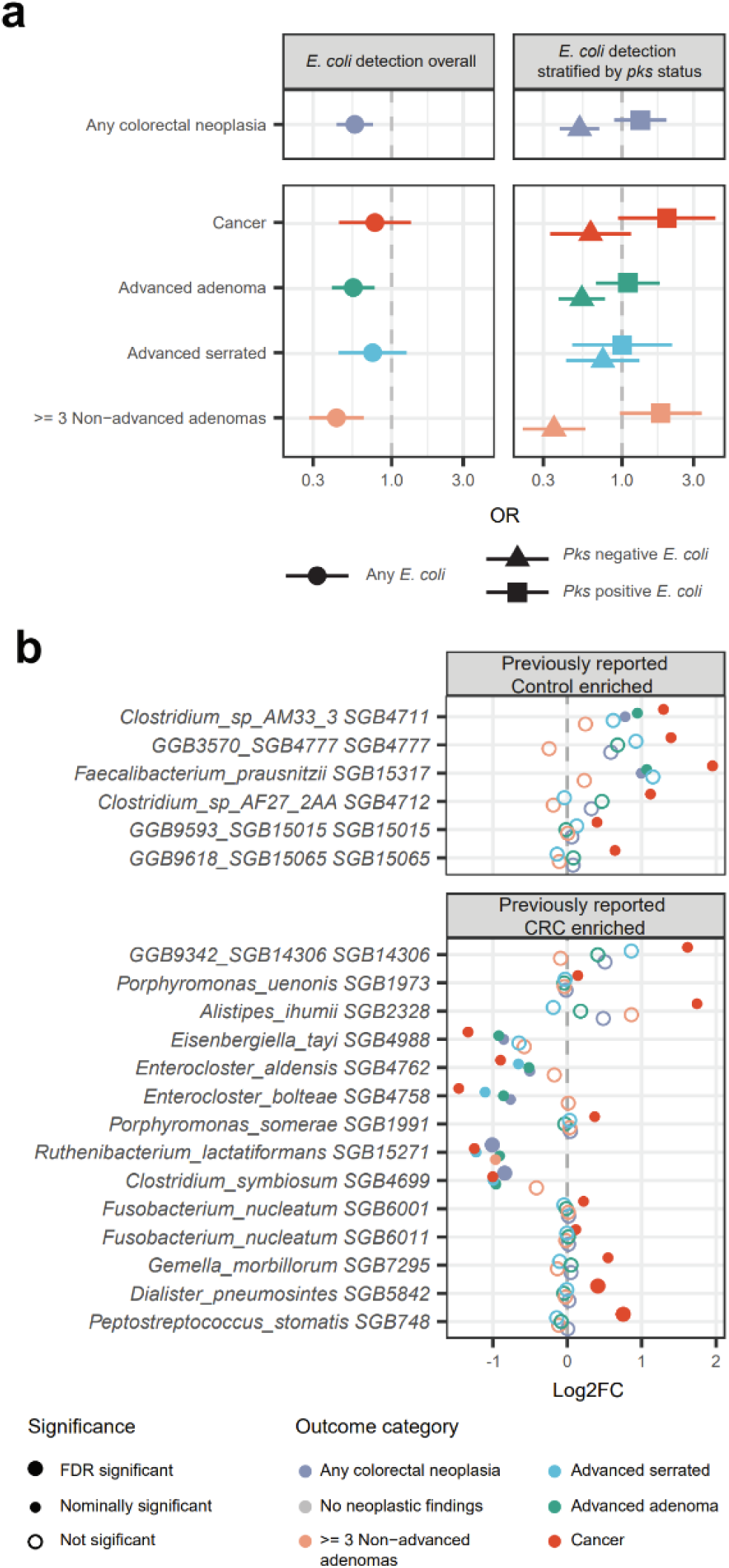
CRC-related depletion of non-pks-encoding E. coli and evaluation of species previously associated with CRC. a) Ranked dot plot displaying the group-wise percentile for each outcome category (x-axis) of each participant for the abundance of E. coli (y-axis). b) Species identified as being associated with the presence or absence of CRC across populations in a large case-control study (n = 122), were evaluated for differences in abundance between those with no neoplastic findings and CRC in the current study. Those with significant differences (p < 0.05) are shown here, where species reported to be more abundant in controls are shown in the top panel, and those reported to be more abundant in CRC are shown in the bottom panel.

Six species were differentially abundant in CRC cases. Four uncharacterized bacteria (one belonging to the genus *Clostridia*, one to the family *Oscillospiraceae*, and two within the phyla Bacillota and Tenericutes) were positively associated with CRC, whereas two *Streptococcus* species, *S. salivarius* and *S. parasanguinis*, were less abundant in CRC cases. Levels of both *S. salivarius* and *S. parasanguinis* were higher in individuals with recent use of proton pump inhibitors (PPIs) or other anti-acid medications (Figure 4c), as has been previously reported^37^. To assess whether the association of these species with CRC was driven by drug use, we conducted a sensitivity analysis stratifying participants by recent antacid/PPI use, showing significant associations also for those without recent use (Supplementary Figure S10). Similarly, differential abundance results in general were consistent for those without recent antibiotics use, and both with and without non-neoplastic findings, although associations were diminished among recent antibiotics users.

Evaluation of 122 species recently identified as associated with CRC^18^, showed that 32% (n = 38) of these had low prevalence (<10%) in our dataset (Supplementary Data 6). Differential abundance analysis including species with at least 2 observations in our dataset resulted in 98/118 showing no evidence of differences in abundance between those with no neoplastic findings and those with CRC. Although nominal significance was observed for 20 species (Figure 5), 11 were enriched in the opposite diagnostic group (no neoplastic findings rather than CRC (n = 5), or vice versa (n = 6); p < 0.05). Species reproducibly enriched in CRC (p < 0.05) included *Peptostreptococcus stomatis*, *Dialister pneumosintes*, *Gemella morbillorum*, *Porphyromonas somerae*, *Alistipes ihumii*, *Fusobacterium nucleatum sensu stricto* (SGB6011), *Fusobacterium nucleatum polymorphum* (SGB6001), *Porphyromonas uesonis* and the uncharacterized bacterium *GGB9342 SGB14306* (belonging to phylum Bacillota), whereas the species *Enterocloster spp. aldensis* and *boltae*, *Eisenbergiella tayi*, *Ruthenibacterium lactatiformans*, and *Clostridium symbiosum*, which were previously reported to be associated with CRC, were here found to be enriched in those with no neoplastic findings.

### Site-specific associations of colorectal lesions and the microbiome

Using detailed information about lesion localization available from endoscopist records, we evaluated differences in the microbiome relative to the localization of advanced lesions in the gut. CRC and advanced adenomas were more commonly detected in recto-sigmoid regions and the cecum, with a lower degree of site-specificity for serrated lesions and non-advanced adenomas (Figure 6a, Supplementary Table S5).

**Figure 6:**
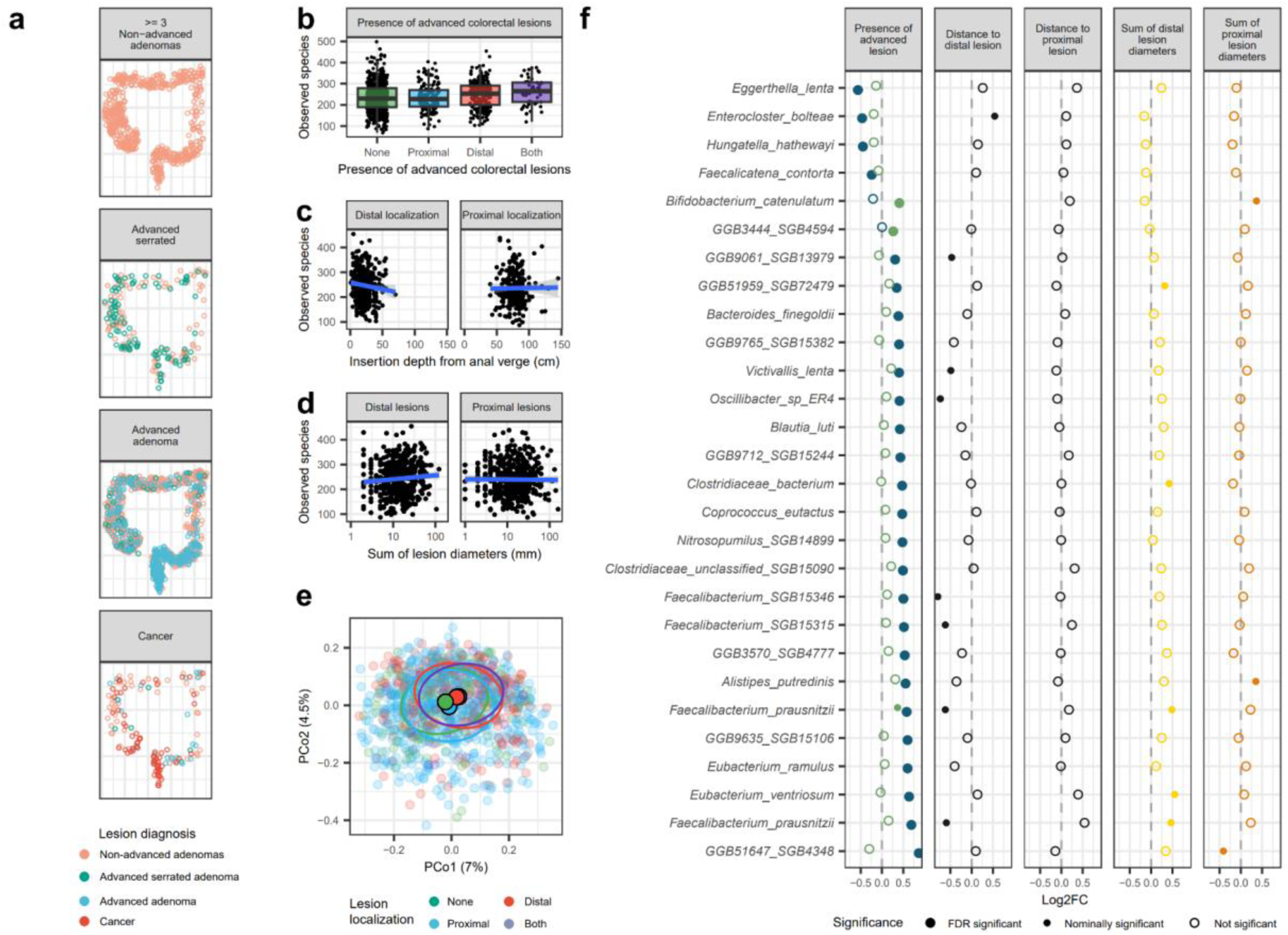
Site-specific association between the microbiome and colorectal lesions. a) Localization of lesions detected at colonoscopy by diagnostic group. Lesions were assessed by a pathologist as part of clinical routine, and if more than one lesion in a patient, the most severe diagnosis was used (see methods). Note that, due to discontinuation of the colonoscopy procedure when CRC was detected, no subsequent lesions (that is, more proximal ones) were registered. b) Number of observed species according to whether advanced colorectal lesions (advanced serrated lesions, advanced adenoma, or CRC) were detected at colonoscopy. Lesions located on the right side or transversal colon up to the splenic flexure, were considered “proximal”, and those distal to this were considered “distal”. с) Species alpha diversity (as in b) by distance from the anal verge to the most severe lesion. The distance was measured using the insertion depth of the colonoscope. Trend lines are based on linear regression with 95% confidence intervals indicated in shaded areas. d) Species alpha diversity and colorectal lesion burden (the sum of lesion diameters for each colonic region). e) PCoA based on Bray-Curtis distances, categorized by findings of advanced colorectal lesions by site (as in b). Solid points indicate centroids, and ellipses encompass 50% of observations. f) Differentially abundant species according to lesion localization and lesion burden according to five alternative models. In panel one, advanced lesions were coded as present or absent, using one variable for distal lesions (blue) and one for proximal lesions (green); in panels two and three, the distance to the most severe lesion was included. In models reported in panels four and five, the localization-specific burden of lesions was included as an independent variable, using separate models for distal and proximal lesions. Filled markers represent significant associations, with marker size denominating nominally (small) and FDR corrected (large) significant results.

Overall, individuals with distal lesions had higher alpha diversity (observed species) (Figure 6b; linear model p < 0.001), which increased the closer lesions were located to the anal verge (Figure 6c; p = 0.04). There was also a positive association between distal lesion burden and species richness (Figure 6d; p = 0.04), strengthening the inference of a localization-dependent association between colorectal lesions and microbial diversity. Regarding proximal lesions, we observed no association between diversity and lesion presence, nor with their distance to the anal verge or lesion burden (Figure 6b-d).

Beta diversity estimates mirrored the alpha diversity findings, with significant differences between distal, but not proximal lesions, when compared to no lesions (Figure 6e; PERMANOVA p = 0.001 for distal, p = 0.10 for proximal). Further, 26 species ere associated with the presence of distal lesions, whereas only 2 were associated with proximal lesions (Figure 6f). Species associated with distal lesions included several of those also associated with neoplastic findings overall (5/13; Supplementary Figure S11), while this was not the case for proximal lesions (0/13).

Interestingly, four out of 12 *Faecalibacterium* species (SGBs) were positively associated with the presence of distal lesions and negatively associated (nominally significant) with the distance of lesions to the anal verge. Two of the *F. prausnitzii* SGBs were also associated with the burden of distal lesions (the sum of lesion diameters; nominal significance). Still, no KOs encoded by members of the *Faecalibacterium* genus were differentially abundant in individuals with distal lesions (Supplementary Table 6).

## Discussion

Despite extensive efforts to characterize the CRC-associated metagenome, few studies have examined screening-relevant populations. In this CRC screening cohort, where inclusion was based on national registry linkage, we employed clinically relevant diagnostic categories and integrated microbiome findings with FIT test values. Our results show that the microbiome of those with colorectal neoplasia differs from those without and contributes to a modest increase in predictive capacity for premalignant lesions compared to the FIT value alone.

Previous studies have achieved high levels of accuracy for the identification of both CRC^15,16,29^ and adenomas^29,38,39^ using microbiome profiling. However, these have often included symptomatic cases or lack well-described participant identification procedures, which, as shown in a recent systematic review, exposes them to a high risk of bias^40^. Evaluating microbiome-based biomarkers for CRC screening requires recruitment strategies that are transparent and minimize bias, but also predictive modeling supporting the dual aims of CRC screening, of both early CRC detection and its prevention through detection of premalignant lesions. Our results show that gut microbiome-based predictions made in a relevant screening setting can improve identification of those who are likely to benefit from colonoscopy, especially for those with premalignant lesions. We addressed the problem of distinguishing neoplasia from other causes of bleeding in FIT-positive participants, most of whom had tested FIT-negative in previous rounds and were asymptomatic, making accurate classification particularly difficult. Additionally, most neoplasms were non-malignant, which has been shown to be harder to identify though analysis of the gut microbiome^41,42^.

The utility of new diagnostic tests depends on how they compare to existing procedures. Participants recruited in the CRCbiome study were FIT-positive at a threshold of 15 μg hemoglobin per gram of feces - a low threshold compared to other screening programs^7,43^, allowing us to compare how the microbiome can contribute to improved prioritization of colonoscopy referrals relative to increases in the FIT value threshold. Here, we show how adding predictions from a microbiome-based model to FIT results could improve classification for premalignant lesions beyond what was achievable by using either the FIT alone or the FIT together with lifestyle and demographic factors. Our results suggest that the microbiome may be a source of biomarkers, with the potential to improve prediction of colonoscopy outcomes in FIT-based screening programs.

In a screening setting, individuals with occult blood in their stools, but without neoplasia, are more likely to have other gastrointestinal conditions or have unhealthy habits associated with an increased rate of intestinal bleeding^44,45^. Several species negatively associated with neoplasia in our study have previously been found to be more abundant in CRC, including *E. coli*, *H. hathewayi*, *E. lenta*, *C. symbiosum*, *E. boltae*, and *F. plautii* either in general^18,46–48^, or as site-^49^, stage-^18^, or age-specific associations^50^. We also found a positive association between microbes that are commonly considered beneficial and the presence of distal lesions, including several *Faecalibacterium* species^51,52^. Our contradictory findings suggest that enrichment of *H. hathewayi*, *E. lenta*, and *C. symbiosum* might be caused by bowel conditions other than CRC development; rather functioning as indicators of an unhealthy gastrointestinal community in general.

We identified *E. coli* as being negatively associated with presence of colorectal neoplasia, and that this association was restricted to *pks*-negative *E. coli*. In contrast, there was a trend towards enrichment of *pks-*positive *E. coli* in individuals with CRC or any colorectal neoplasia. The genotoxic effect of colibactin produced by *pks*-positive *E. coli* has been demonstrated^11^, with mutations caused by colibactin likely being an early event in carcinogenesis^53,54^. While a recent large study of Dutch screening participants cast doubt on the utility of *pks*-positivity alone as a marker in CRC screening^55^, our results suggest that FIT-positivity should also be considered when evaluating the role of *pks*-positivity for *E. coli* in CRC screening. Supporting such a conclusion, Chénard et al., while not stratifying by *pks* status, showed that *E. coli* was overrepresented among neoplasia-negative screening participants with fecal occult blood^29^, although such enrichment was not reported in another study^56^.

The enrichment of oral bacteria in the gut microbiome of individuals with CRC has been shown in multiple reports^15,18,39,57^. We reproduce several of the most consistently reported associations in this screening population, including enrichment of *F. nucleatum* and *P. stomatis*. Although the prevalence of these species was limited, our results confirm these species to be specifically enriched in CRC. We also found a depletion of some oral bacteria in those with CRC, including several *Streptococcus* species. The *Streptococcus* genus comprises both pathogenic, commensal, and probiotic species and strains. The association of *Streptococcus* species with CRC is also mixed: while bacteremia with *S. gallolyticus* (previously known as *S. bovis)* has a long-established link to the presence of CRC^58^, other members of the genus, including *S. salivarius*, have been found to be depleted in CRC^59^. Interestingly, *S. salivarius*, which was among the CRC-depleted streptococci in our study, has been shown to inhibit the growth of *F. nucleatum* through secretion of nisins^60^, with further evidence for a protective effect for other *Streptococcus* species^61^. While the abundance of several *Streptococcus* species has previously been associated with PPI use^37,62–64^, we found that the depletion of streptococci in CRC was independent of recent anti-acid medication use.

Overall characteristics of the microbiome were associated with CRC-related outcomes. Our classification into four community types, dominated by e.g., *P. copri* and *Bacteroides* species, is consistent with other reports^28,34,64^. We found community states to have remarkably different associations with outcome groups, where a *Bacteroides* spp. enriched state was strongly associated with a lack of neoplasia. Moreover, prediction accuracy for colorectal neoplasia differed between the microbial community types, suggesting that such community types could serve as a basis for tailored screening. Still, larger studies are needed to validate such an approach.

Our results showed microbial diversity to be elevated among those with advanced distal lesions. This is in line with results from a recent study also identifying elevated diversity in distal CRC compared to proximal cases and healthy controls^18^. Additionally, we found a novel increase in diversity associated with both distal lesion proximity to the anal verge and distal lesion load, providing further support to the conclusion that distal lesions are associated with increased diversity. Similarly, we found most differentially abundant species to align with the presence or absence of distal lesions, but not of proximal ones. These findings could indicate that lesion-associated microbial alterations are more readily measured distally, likely due to dilution or degradation of signals during colonic transit, or alternatively, reflecting cellular differences between distal and proximal lesions.

Recent reports have highlighted the importance of confounding in CRC-microbe associations^28^. We observed several cases where associations come to light only after making statistical adjustments. As such, depletion of *F. plautii* in advanced serrated lesions became evident only after statistical adjustments for lifestyle and demographic factors.

This study has notable strengths. We employed standardized participant recruitment strategies, sample collection procedures, and laboratory protocols to ensure uniform capture of microbiome signatures representative of a colorectal screening population. Study participants were deeply characterized using validated questionnaire data and complete registry-based prescription drug use histories. Outcome measures were based on detailed clinicopathological records from centralized screening centers. There are also several important limitations. Using surplus FIT buffer with low stool concentration likely limited the sensitivity for very low-abundance species. Further, the study design exclusively included FIT-positive participants, limiting the generalizability of our findings. Lastly, basing recruitment on consecutive screening cases resulted in few CRC cases, likely causing CRC-directed models to be underpowered.

Even modest improvements in the accuracy of screening models can save lives and reduce the burden on healthcare systems. Especially in settings where colonoscopy resources are limited, a more accurate pre-screening tool may improve CRC screening by making it more efficient. By combining microbiome-based models with the FIT test and lifestyle exposure, we could distinguish those with a need for colonoscopy due to the presence of premalignant lesions or CRC more accurately than FIT alone. These results indicate that the microbiome may be used to improve screening-based early detection – which is precisely where the potential for improvement is greatest. Still, refinement of markers and external validation is warranted.

## Conclusion

We show that the gut microbiome can contribute to more accurate classification of colorectal neoplasia in a CRC screening population than FIT alone, highlighting the potential of microbial biomarkers to improve early detection when combined with FIT. Our findings reveal specific microbial signals associated with CRC screening outcomes. We reproduce known CRC-associated enrichments of bacteria such as *F. nucleatum* and *P. stomatis* and show that *pks*-negative *E. coli* is enriched in participants without neoplastic findings. In contrast, bacteria such as *F. prausnitzii*, *H. hathewayi*, *E. lenta*, and *C. symbiosum* appear to be associated with other conditions causing FIT positivity rather than with CRC development. Both overall microbial community structure and specific taxa were associated with the presence of distal lesions, while associations with proximal lesions were limited.

## Methods

The CRCbiome project complies with all relevant ethical regulations, and all participants provided informed consent. The CRCbiome project has been approved by the Regional Committees for Medical Research Ethics South East Norway (REK approval no. 63148).

### Subject recruitment and sample collection

Participants in the CRCbiome study were recruited from FIT-positive participants in the Bowel Cancer Screening in Norway (BCSN) study, a randomized controlled trial (ClinicalTrials.gov: NCT01538550) comparing the effectiveness of once-only sigmoidoscopy and biennial FITs, testing for fecal occult blood^31^. In the FIT arm of BCSN, men and women in the age range 50–74 years in 2012 (born between Jan 1, 1938, and Dec 31, 1962), residing in one of two closely situated, but geographically distinct regions in South-East Norway (Moss (region 1), or Bærum (region 2)), were invited to self-collect stool samples using provided sampling kits suitable for FIT analysis, with no dietary or medication restrictions prior to sampling. Participants shipped stool samples by mail to a central laboratory for FIT testing (estimated 3–10 days shipping time). Hemoglobin concentration (FIT-value) was measured using the OC-Sensor Diana system (Eiken Chemical, Tokyo, Japan), and the sampling tube with remaining buffer (containing about 10 mg feces) was stored at −80 °C. Those with a positive FIT result (>15 µg hemoglobin/g feces, i.e. fecal occult blood detected) were scheduled for a follow-up colonoscopy, and further FIT screening rounds were discontinued.

BCSN participants who tested FIT positive between October 2017 to March 2021 were invited to take part in CRCbiome. Invitation letters were sent two to 14 days after FIT test results with invitations to follow-up colonoscopy. CRCbiome recruited men and women partaking in their second, third, or fourth scheduled round of FIT screening as part of the BCSN trial. Out of 2700 individuals invited to CRCbiome, 1640 provided informed consent. Further details on recruitment procedures can be found in Kværner *et al*. ^32^.

### Clinical examination, diagnosis and outcome definitions

After a FIT-positive screening result, participants were scheduled for colonoscopy at a screening center (one for each region) preceded by a bowel cleansing procedure. During colonoscopy, localized lesions (mostly polypous lesions) were removed and submitted for histopathological examination. Participants with no findings during colonoscopy were categorized as “negative” based on the endoscopist’s assessment. All other lesions were classified by a pathologist according to standard clinical routine and resulted in categories based on their most severe finding ranging from “non-neoplastic lesions”, “other findings”, “non-advanced serrated polyps”, “1-2 non-advanced adenomas”, “3 or more non-advanced adenomas”, “advanced serrated lesion”, “advanced adenoma” and “cancer”. Those with non-neoplastic findings included endoscopy-detected presence of hemorrhoids, IBD, diverticulitis, or other conditions not related to CRC, and those with other findings included inflammatory polyps and granulated tissue polyps. As we considered those with non-advanced serrated lesions and 1 or 2 non-advanced adenomas to be a low-risk group in terms of future development of CRC, this group was not included in the metagenome analyses seeking to evaluate the gut microbiome as a source of biomarkers for CRC screening.

CRC was defined as adenocarcinoma of the colon or rectum (ICD10 codes C18-20), and advanced adenomas were defined as adenomas ≥10 mm, or those with ≥25% villous histology, or with high-grade dysplasia. Advanced serrated lesions included any serrated lesions (hyperplastic polyp, sessile serrated lesion, or traditional serrated adenoma) with a size of ≥10 mm or dysplasia.

Lesion localization for all lesions detected during colonoscopy was recorded by the endoscopist and categorized according to colonic region as proximal when it was located proximally to, or in the splenic flexure, or as distal. Due to discontinuation of colonoscopy upon detection of CRC, subsequent lesions (more proximal ones) were commonly not registered. Lesion location was also registered as its distance from the anal verge, measured by insertion depth of the colonoscope. For each participant, the distance from the anal verge to the most severe lesion was recorded. As a measure of lesion burden, we calculated the sum of lesion diameters. The latter two measures were calculated separately for the proximal and distal regions of the colon.

### DNA extraction, library generation and metagenome sequencing

FIT cartridges were retrieved and thawed, and leftover buffer was used as input material for DNA extraction and shotgun metagenome sequencing library preparation, as previously described^32^. Briefly, 500 µl aliquots (approximately 30% of remaining buffer) were used for DNA extraction using the QIAsymphony DSP Virus/Pathogen Midikit (Qiagen), with offboard lysis protocol based on bead-beating. Extracted DNA was eluted in 60 µl AVE buffer (Qiagen). For samples with a DNA concentration of less than 1.5 ng/µl, DNA extraction was repeated using a second aliquot. Screening samples with a DNA concentration of 0.7 ng/µl or more were considered eligible for metagenome library preparation. Sequencing libraries were generated according to the Nextera DNA Flex Library Prep Reference Guide, with the exception of scaling down the reaction volumes to ¼ of the reference. Library pools of 240 samples were combined and size-selected to a fragment size of 650–900 bp. Sequencing was performed using the Illumina NovaSeq system using S4 flow cells with lane divider, with each pool sequenced on a single lane resulting in paired-end 2×151 bp reads. Sequencing output has been reported previously^65^.

### Taxonomic classification, determination of functional potential, and establishment of metagenome-assembled genomes (MAGs)

Sequencing reads were processed for removal of adapters, low-quality bases, and reads mapping to the human genome using kneaddata (v0.12) with standard settings. Read-based taxonomy and gene content were assessed using MetaPhlAn4 (v4.0.6) and HumanN3 (v3.7)^66,67^, respectively, with the mpa_vOct22_ChocoPhlAn_202212 pangenome database, using the UniRef90 database to assign gene families to KEGG orthogroups (KOs). Read-based taxonomic abundance was evaluated at the level of species-level genome bins (SGBs; referred to as “species” in the text) using relative abundance determined by MetaPhlAn4. Gene content abundance derived from HumanN3 analysis was scaled by the number of quality-controlled reads per million.

Separately, the metagenome processing framework Metagenome-ATLAS^68^ (v2.4.3) was used for sequencing QC, assembly, binning, generation of representative genomes (MAGs), and cataloging of gene content. In brief, low-quality bases and reads were filtered, and human and phiX sequences were removed using BBMap (v37.99)^69^. Remaining reads passing QC were assembled using MetaSpades^70^ (v3.13) with default settings. Assemblies were grouped into genomes using DAStool^71^ (v1.1), based on genomic bins identified by MetaBat^72^ (v2.2) and MaxBin^73^ (v.2.14). Representative genomes were defined by dereplication using dRep^74^ (v2.2) with a 95% sequence identity for 60% genome overlap. Only genomes with a quality score Q ≥ 50 were retained, where the quality score was defined based on CheckM metrics as Q = completeness-5×contamination. Taxonomy was assigned using the GTDB-Tk^75^ (v1.3), with the GTDB database (v95). The abundance of MAGs was determined by calculating the median depth of reads mapping to 1000-bp bins of each genome and was scaled by the number of reads per million. For evaluation of consistency between read-based (MetaPhlAn4) and assembly-based taxonomic characterization, we matched species names as assigned using GTDB annotations for both datasets. Species uniquely identified in both datasets were compared across samples by prevalence in each dataset, and the cosine similarity measure of their abundance across samples.

A publicly available dataset of gut metagenomes^34^ derived from conventionally collected stool samples was retrieved and subjected to characterization of species abundance using MetaPhlAn4 using the procedure described for CRCbiome. Comparisons were made of mean relative abundance for each species and species-based beta-diversity using a principal coordinates analysis (PCoA) visualization.

### Evaluation of microbial diversity and community types

Microbial alpha diversity was measured as the number of observed species (or KOs in the case of gene data), and by the Shannon and inverse Simpson diversity indices based on relative abundance of species or KOs and used in analyses as continuous variables. Species- and KO-based beta diversity was calculated using the Bray-Curtis dissimilarity measure.

Microbial community types were defined using Dirichlet Multinomial Mixture^35^ (DMMs) models as implemented in the R package ‘DirichletMultinomial’, based on the relative abundance of species with a minimum abundance of 0.1% in 10% of individuals. The number of community states (components; k) was determined by selecting the number of components from 1-8, resulting in the lowest Laplace approximation score. Species contribution to the community type definition was defined as the difference in contribution between a single-component model and each community of a four-component model.

### Detection of *pks*-positive *E. coli*

Presence of the *pks* gene cluster in *E. coli* genomes was determined by mapping reads to the pks gene cluster (Genbank AM229678.1). Here, any individual with *E. coli* detected using MetaPhlAn4 and with reads covering at least 10% of the *pks* module was classified as *pks* positive, unless read mapping showed the colibactin gene cluster to be incomplete.

### Questionnaire data

Data on regular diet, lifestyle factors, and demography were collected using a lifestyle and demography questionnaire (LDQ) and a validated food frequency questionnaire (FFQ) designed to register the habitual diet over the last year. Details on the two questionnaires have been described previously^32^. For the FFQ, those with low-quality questionnaires (two or more missing pages, too many missing food items and/or portion sizes, or inconsistent reporting; n = 21), too low energy intake (<600 and <800 kcal/day for women and men, respectively; n = 11), or too high energy intake (>3500 and >4200 kcal/day for women and men, respectively; n = 46) were excluded. No exclusion criteria were applied for the LDQ, and missing data was handled on a question-by-question basis.

The questionnaires provided information on factors with strong evidence for an increased risk (consumption of red and processed meat and alcohol, body fatness, adult attained height, and smoking) or a decreased risk of CRC (intake of dairy products, calcium, wholegrains, and fiber, and level of physical activity). It further provided information on the consumption of main food groups (fruit and berries, vegetables, fish, and sugar-sweetened beverages), as well as total energy intake, being positively correlated to many of the included foods and nutrients^76^.

Variables derived from questionnaires that were used in statistical models (see below) included smoking status, level of education, recent antibiotic use, and a composite score defining adherence to cancer prevention recommendations. Smoking status was categorized as smoker or non-smoker, where smokers included current smokers (any level), or those who reported having ceased smoking within 10 years, or who had smoked for 30 years. Non-smokers included never-smokers and current non-smokers who ceased smoking at least 10 years prior to study inclusion and reported having smoked for less than 30 years. Education level was recorded as the highest level of completed education (primary school, high school, or college/university). Recent antibiotic use was defined using self-reported use and registry data (see below). Based on the questionnaire data, we composed a seven-point score measuring adherence to the recommendations in the 2018 World Cancer Research Fund/American Institute for Cancer Research (WCRF/AICR) Cancer Prevention Report(referred to as the WCRF score, operationalization described in a separate paper^21^). The score measured adherence to the following recommendations: 1) be a healthy weight, 2) be physically active, 3) consume a diet rich in wholegrains, vegetables, fruits, and beans, 4) limit consumption of “fast foods” or other processed foods high in fats, starches, and sugars, 5) limit consumption of red and processed meat, 6) limit consumption of sugar sweetened drinks, and 7) limit alcohol consumption. Adherence to each recommendation was assigned a score of 0, 0.5, or 1, with increasing degree of adherence; the WCRF score was defined as the sum of these scores. The 2018 WCRF Score has been linked to colorectal carcinogenesis in several longitudinal studies^77–80^, including the current study population^21^. A complete list of the diet, lifestyle, and demography variables employed in the study can be found in Supplementary Data 7.

### Self-reported medication use and prescription drug dispensation

The LDQ included questions about use of antibiotics or anti-acid drugs (antacids or proton pump inhibitors (PPIs)) within 3 months of answering the questionnaire. For each participant, we linked records of dispensed prescription drugs provided by the Norwegian Prescription Database (NorPD), which maintains records of dispensed prescription drugs in Norway, classified according to the Anatomical Therapeutic Chemical (ATC) system^81^. Drugs that are purchased without prescriptions or are supplied to hospitals or nursing homes are not included. For antibiotics and anti-acid drug use, recent use was recorded if participants had self-reported use, or registry data showed dispensation of drugs of the same classes (ATC codes J01, A07AA, or P01AB for antibiotics, and A02A or A02B for antacids/PPIs) within 4 months.

Data from NorPD was also used for evaluation of study population representativeness. Here, data included NorPD records for CRCbiome participants, as well as for two comparison populations: 1) the remaining participants of the FIT arm of the BCSN trial, and 2) a nationally representative population: a random draw of 16 500 Norwegians eligible for bowel screening in 2017 (born in 1946-1962). We obtained information on dispensation of prescription drugs, indicating common comorbidities, or that were of relevance to microbiome analyses. These included drugs for diabetes (≥2 dispensations of any drug with ATC code A10); cardiovascular disease (≥1 of any with ATC codes C, and/or B01); chronic obstructive pulmonary disease (≥1 of any with ATC codes R03AC, R03AK, R03AL, R03BB, R03DA, R03DC, R03DX, R03BA); and antibiotics (≥1 of any with ATC codes J01, A07AA, or P01AB). Prescription drug dispensation was assessed within 12 and 24 months prior to participant inclusion. For BCSN participants, inclusion was defined as the date of invitation to the first round of biennial screening. For the population controls, a fictitious inclusion date was set as a random date in 2017. For CRCbiome participants, two definitions of inclusion were used: 1) the invitation date for the first round of biennial screening (i.e., their inclusion to BCSN), and 2) the invitation date for the FIT screening round in which they tested positive and were invited to CRCbiome.

### Statistical analysis

For descriptive statistics, categorical variables were summarized as counts and row-wise percentage, and continuous variables as medians with interquartile range (Q1, Q3). Associations between microbial variables and participant characteristics were evaluated in models adjusted for age, sex and region, and continuous variables were encoded as tertiles (low, mid, and high), where the lower level was treated as reference. Results of associations for the groups “unknown”, “missing”, or those with less than 10 observations were included in statistical analyses but not reported separately. Evaluation of associations between microbial variables and outcomes (any colorectal neoplasia, diagnostic groups, or localization-dependent measures of colorectal lesions) were modeled with adjustments for age, sex, region, the WCRF score, smoking status, level of education, findings of hemorrhoids, findings of IBD, and recent antibiotics use unless noted otherwise. Participant age and the WCRF score were included as continuous variables, whereas sex, region, smoking status, level of education, findings of hemorrhoids and IBD, and recent antibiotics use were included as categorical variables. The selection of covariates was largely based on the latest Expert Reports from WCRF/AICR of 2018, summarizing the available evidence on diet, physical activity, body weight, and CRC risk^19,82^.

Assessment of associations with site-specific lesions was performed in separate models for each measure, employing the presence, burden, or distance to the anal verge of lesions as independent variables. For the presence of advanced distal and proximal lesions, one dummy variable was included as an independent variable for distal lesions, and one for proximal lesions. Modeling of associations with the distance to colorectal lesions, or of the burden of colorectal lesions, only included participants with lesions, and for models of one colorectal region, those with lesions in the other region were excluded.

Models including alpha diversity were also adjusted for sequencing depth. Differences in alpha diversity between groups of participants defined by host characteristics were assessed using linear regression models, with alpha diversity as the dependent variable. Differences in alpha diversity or other microbially defined groups (community states, *pks*-stratified *E. coli*) related to outcome categories were assessed using multinomial logistic regression models with outcome as the dependent variable, the standard adjustment variables.

Differences in beta diversity between groups were evaluated using permutational analysis of variance (PERMANOVA) as implemented in the R package vegan^83^ using the “adonis2” function with 999 permutations. For evaluation of differences between levels of categorical variables, pairwise tests were performed. PERMANOVA tests were run using marginal testing, with adjustment for age, sex, and region.

### Machine learning/classification

Machine learning for classification of participants by the presence of colorectal neoplasia was carried out using three categories of datasets: 1) species abundance estimated using either read-based taxonomic profiling or MAGs; 2) abundance of KEGG orthologous (KO) groups using read-based (HumanN3) quantification; or 3) a set of diet, lifestyle, and demographic factors capturing broad features of demography and participant lifestyle (Supplementary Data 7).

Models were trained and evaluated using the tidymodels^84^ framework with a 20-fold repeated 5-fold cross-validation (CV) within an 80% training set, with final models additionally evaluated in a 20% test set. The test set was randomly selected, being matched with the training set by age, sex, and screening result. Algorithm selection, parameter tuning, dataset specifications, and participant stratifications were explored exclusively within the training dataset. Evaluations of model performance in assigning class probabilities were made by assessing the mean area under the receiver operating characteristic curve (AUROC) across CV partitions. Features with low prevalence (<70%) or with low degree of variation (the most common value 19 times more frequent than the second most common) were excluded from prediction models. For unbalanced datasets (one outcome category constituting more than 2/3 of the dataset), the adaptive synthetic algorithm^85^ was used for upsampling the minority group.

Ensemble models were set up using stacking with logistic regression models including predictions made with microbial models and a set of demographic and clinical variables, including 1) log-transformed FIT values, and 2) log-transformed FIT values, sex, age, and the WCRF score. Additionally, logistic regression models were set up for the latter set of variables without the microbial predictions. Ensemble models were trained on datasets limited to those without neoplastic lesions or with advanced adenomas or CRC.

### Differential abundance analyses

Differential abundance was assessed using Microbiome Multivariable Associations with Linear Models (MaAsLin2)^86^. To account for differences in KO abundance largely explained by taxonomy, KOs with high correlation to at least one species (Pearson’s product moment correlation coefficient above 0.5) were excluded. Further, only features present in at least 10% of samples were retained for analyses. Analyses were performed using a linear model with log transformation and total sum scaling normalization. Differential abundance analyses evaluating associations with outcome categories were adjusted for age, sex, region, sequencing depth, smoking status, level of education, the WCRF score, hemorrhoids, IBD, and recent use of antibiotics. Sensitivity analyses were performed, stratifying by recent use of antibiotics (see definition above), recent use of anti-acid medication, or the detection of any non-neoplastic lesions during colonoscopy.

Characterization of host-microbe associations was carried out using MaAsLin2 analyses contrasting levels of host characteristics (as described for alpha- and beta-diversity), adjusting for age, sex, region, and sequencing depth, with results reported for species with significant outcome associations.

To evaluate associations reported in the literature, we extracted a list of microbial species identified as being associated with CRC or controls across populations in a recent study^18^. We restricted the set of microbes to be evaluated to those that were significant in their models adjusting for sex, age, and BMI (FDR < 0.05), which included 122 SGBs. For evaluation of these species, we carried out a differential abundance analysis using MaAsLin2, including those observed in at least two individuals and considered statistical significance without FDR correction.

To evaluate *Faecalibacterium* functional potential, we extracted from species resolution HumanN3 data all KOs annotated as belonging to *Faecalibacterium* species KOs and normalized read counts per 1 million reads. Differential abundance for each of these KOs was assessed using a Kruskal-Wallis test with FDR correction.

## Acknowledgements

We would like to express our gratitude to Jan Inge Nordby from the Department of Medical Biochemistry at Oslo University Hospital, and Erik Natvig and Anita Jørgensen from the Department of Colorectal Cancer Screening at the Cancer Registry of Norway (CRN), National Institute of Public Health (NIPH) for their contributions to biobanking, laboratory work, and data management. We also wish to thank Harri Kangas and Pekka Ellonen at the Finnish Institute of Molecular Medicine (FIMM) for their assistance in preparing sequencing data; library preparation and sequencing were conducted at the FIMM Technology Centre, supported by HiLIFE and Biocenter Finland. Our thanks extend to Martin Solbrekken and Stine Langbråten at the CRN, NIPH, along with Anne Marte Wetting Johansen at Department of Nutrition, University of Oslo (UiO), for their essential contributions to scanning and processing the questionnaire data. Additionally, we acknowledge the support of current and former group members for their administrative and scientific contributions, including Elina Vinberg, Maja Sigerseth Jacobsen, Even Sannes Riiser, Jonas Thy, Astrid Riseth Andersen, Paula Istvan, and Lisi Zhan. Research reported in this publication was supported by the Norwegian Cancer Society, projects 190179 (TBR) and 198048 (PB), and the South-East Norway Regional Health Authority projects 2022067 (TBR) and 2020056 (TBR).

## Data availability

DNA sequencing data generated in this study are deposited in Federated EGA under accession code EGAS50000000170 (https://ega-archive.org/studies/EGAS50000000170). Per participant consent, submitted FASTQ files exclude reads mapping to the human genome. Processing of data from this study must comply with the General Data Protection Regulation (GDPR). Access can be obtained by following the procedure described here: https://www.mn.uio.no/bils/english/groups/rounge-group/crcbiome/. All scripts are available at https://github.com/Rounge-lab/Microbiome-signatures-CRCbiome.

## Ethics declarations

### Conflict of interest declaration

The bowel preparation used for colonoscopy in the BCSN trial was provided free of charge by Ferring Pharmaceuticals; the company had no role in study design, data collection and analysis, decision to publish or preparation of the manuscript. The authors report no other conflicts of interest.

## Abbreviations

AICR: American Institute for Cancer Research
AUROC: Area Under the Receiver Operating Characteristic Curve
ATC: Anatomical Therapeutic Chemical
BCSN: Bowel Cancer Screening in Norway
BMI: Body mass index
CI: Confidence interval
CV: Cross-validation
CRC: Colorectal cancer
FFQ: Food frequency questionnaire
FIT: Fecal immunochemical test
IBD: Inflammatory Bowel Disease
KEGG: Kyoto Encyclopedia of Genes and Genomes
KO: KEGG orthogroups
LDQ: Lifestyle and demography questionnaire
MaAsLin: Microbiome multivariable associations with linear models
MAG: Metagenome-assembled genome
PCoA: Principal coordinate analysis
PERMANOVA: Permutational multivariate analysis of variance
PPI: Proton Pump Inhibitor
RF: Random Forest
SD: Standard deviation
WCRF: World Cancer Research Fund

## References

1. Schreuders, E. H. et al. Colorectal cancer screening: a global overview of existing programmes. Gut 64, 1637–1649 (2015).

2. Shaukat, A. & Levin, T. R. Current and future colorectal cancer screening strategies. Nat Rev Gastroenterol Hepatol 19, 521–531 (2022).

3. Doubeni, C. A. et al. Fecal Immunochemical Test Screening and Risk of Colorectal Cancer Death. JAMA Network Open 7, e2423671 (2024).

4. Imperiale, T. F., Gruber, R. N., Stump, T. E., Emmett, T. W. & Monahan, P. O. Performance Characteristics of Fecal Immunochemical Tests for Colorectal Cancer and Advanced Adenomatous Polyps: A Systematic Review and Meta-analysis. Ann Intern Med 170, 319– 329 (2019).

5. Kim, N. H. et al. Are Hemorrhoids Associated with False-Positive Fecal Immunochemical Test Results? Yonsei Medical Journal 58, 150–157 (2017).

6. Therkildsen, S. B., Larsen, P. T. & Njor, S. H. Screening participants with inflammatory bowel disease or high colorectal cancer risk in Denmark: a cohort study. J Public Health Pol 45, 727–739 (2024).

7. Randel, K. R. et al. Performance of Faecal Immunochemical Testing for Colorectal Cancer Screening at Varying Positivity Thresholds. Alimentary Pharmacology & Therapeutics 61, 122–131 (2025).

8. Thomsen, M. K. et al. Risk-stratified selection to colonoscopy in FIT colorectal cancer screening: development and temporal validation of a prediction model. Br J Cancer 126, 1229–1235 (2022).

9. Wong, S. H. et al. Gavage of Fecal Samples From Patients With Colorectal Cancer Promotes Intestinal Carcinogenesis in Germ-Free and Conventional Mice. Gastroenterology 153, 1621–1633 e6 (2017).

10. Cao, Y. et al. Long-term use of antibiotics and risk of colorectal adenoma. Gut 67, 672–678 (2018).

11. Pleguezuelos-Manzano, C. et al. Mutational signature in colorectal cancer caused by genotoxic pks(+) E. coli. Nature 580, 269–273 (2020).

12. Castellarin, M. et al. Fusobacterium nucleatum infection is prevalent in human colorectal carcinoma. Genome Res 22, 299–306 (2012).

13. Kostic, A. D. et al. Genomic analysis identifies association of Fusobacterium with colorectal carcinoma. Genome Res 22, 292–8 (2012).

14. Senthakumaran, T. et al. Microbial dynamics with CRC progression: a study of the mucosal microbiota at multiple sites in cancers, adenomatous polyps, and healthy controls. Eur J Clin Microbiol Infect Dis 42, 305–322 (2023).

15. Thomas, A. M. et al. Metagenomic analysis of colorectal cancer datasets identifies cross-cohort microbial diagnostic signatures and a link with choline degradation. Nat Med 25, 667–678 (2019).

16. Wirbel, J. et al. Meta-analysis of fecal metagenomes reveals global microbial signatures that are specific for colorectal cancer. Nat Med 25, 679–689 (2019).

17. Yachida, S. et al. Metagenomic and metabolomic analyses reveal distinct stage-specific phenotypes of the gut microbiota in colorectal cancer. Nat Med 25, 968–976 (2019).

18. Piccinno, G. et al. Pooled analysis of 3,741 stool metagenomes from 18 cohorts for cross-stage and strain-level reproducible microbial biomarkers of colorectal cancer. Nat Med 31, 2416–2429 (2025).

19. World Cancer Research Fund/American Institute for Cancer Research. Continuous Update Project Expert Report 2018. Diet, Nutrition, Physical Activity and Colorectal Cancer. Available at Dietandcancerreport.Org. (2017).

20. Kværner, A. S. et al. Associations of red and processed meat intake with screen-detected colorectal lesions. British Journal of Nutrition 129, 2122–2132 (2023).

21. Kværner, A. S. et al. Associations of the 2018 World Cancer Research Fund/American Institute of Cancer Research (WCRF/AICR) cancer prevention recommendations with stages of colorectal carcinogenesis. Cancer Medicine 12, 14806–14819 (2023).

22. Kværner, A. S. et al. The association between alcohol consumption and colorectal carcinogenesis is partially mediated by the gut microbiome. MedRxiv (preprint) 2024.10.17.24315656 (2024) doi:10.1101/2024.10.17.24315656.

23. Asnicar, F. et al. Microbiome connections with host metabolism and habitual diet from 1,098 deeply phenotyped individuals. Nat Med 27, 321–332 (2021).

24. Walker, R. L. et al. Population study of the gut microbiome: associations with diet, lifestyle, and cardiometabolic disease. Genome Medicine 13, 188 (2021).

25. Falony, G. et al. Population-level analysis of gut microbiome variation. Science 352, 560–4 (2016).

26. Bucher-Johannessen, C. et al. Women and men exhibit distinct gut microbial profiles linked to colorectal cancer development. 2025.05.16.25327767 Preprint at 10.1101/2025.05.16.25327767 (2025).

27. Wirbel, J., Essex, M., Forslund, S. K. & Zeller, G. A realistic benchmark for differential abundance testing and confounder adjustment in human microbiome studies. Genome Biol 25, 247 (2024).

28. Tito, R. Y. et al. Microbiome confounders and quantitative profiling challenge predicted microbial targets in colorectal cancer development. Nat Med 30, 1339–1348 (2024).

29. Young, C. et al. Microbiome Analysis of More Than 2,000 NHS Bowel Cancer Screening Programme Samples Shows the Potential to Improve Screening Accuracy. Clin Cancer Res 27, 2246–2254 (2021).

30. Feng, Q. et al. Gut microbiome development along the colorectal adenoma–carcinoma sequence. Nat Commun 6, 6528 (2015).

31. Randel, K. R. et al. Colorectal Cancer Screening With Repeated Fecal Immunochemical Test Versus Sigmoidoscopy: Baseline Results From a Randomized Trial. Gastroenterology 160, 1085–1096.e5 (2021).

32. Kvaerner, A. S. et al. The CRCbiome study: a large prospective cohort study examining the role of lifestyle and the gut microbiome in colorectal cancer screening participants. BMC Cancer 21, 930 (2021).

33. Dekker, E., Tanis, P. J., Vleugels, J. L. A., Kasi, P. M. & Wallace, M. B. Colorectal cancer. Lancet 394, 1467–1480 (2019).

34. Lee, J. W. J. et al. Association of distinct microbial signatures with premalignant colorectal adenomas. Cell Host & Microbe 31, 827–838.e3 (2023).

35. Holmes, I., Harris, K. & Quince, C. Dirichlet Multinomial Mixtures: Generative Models for Microbial Metagenomics. PLOS ONE 7, e30126 (2012).

36. Arthur, J. C. et al. Intestinal Inflammation Targets Cancer-Inducing Activity of the Microbiota. Science 338, 120–123 (2012).

37. Aasmets, O. et al. A hidden confounder for microbiome studies: medications used years before sample collection. mSystems 0, e00541–25 (2025).

38. Wu, Y. et al. Identification of microbial markers across populations in early detection of colorectal cancer. Nat Commun 12, 3063 (2021).

39. Baxter, N. T., Ruffin, M. T. th, Rogers, M. A. & Schloss, P. D. Microbiota-based model improves the sensitivity of fecal immunochemical test for detecting colonic lesions. Genome Med 8, 37 (2016).

40. Manning, S. et al. Systematic Review: The Relationship Between the Faecal Microbiome and Colorectal Neoplasia in Shotgun Metagenomic Studies. Alimentary Pharmacology & Therapeutics 62, 568–584 (2025).

41. Goedert, J. J. et al. Reusing a prepaid health plan’s fecal immunochemical tests for microbiome associations with colorectal adenoma. Sci Rep 12, 14801 (2022).

42. Obón-Santacana, M. et al. Meta-Analysis and Validation of a Colorectal Cancer Risk Prediction Model Using Deep Sequenced Fecal Metagenomes. Cancers 14, 4214 (2022).

43. Navarro, M., Nicolas, A., Ferrandez, A. & Lanas, A. Colorectal cancer population screening programs worldwide in 2016: An update. WJG 23, 3632 (2017).

44. Wang, S.-Y. et al. Factors associated with false fecal immunochemical test results in colorectal cancer screening. World Journal of Gastrointestinal Oncology 17, 101487 (2025).

45. Amitay, E. L., Cuk, K., Niedermaier, T., Weigl, K. & Brenner, H. Factors associated with false-positive fecal immunochemical tests in a large German colorectal cancer screening study. International Journal of Cancer 144, 2419–2427 (2019).

46. Qin, Y. et al. Consistent signatures in the human gut microbiome of old- and young-onset colorectal cancer. Nat Commun 15, 3396 (2024).

47. Zhang, J. et al. Expansion of Colorectal Cancer Biomarkers Based on Gut Bacteria and Viruses. Cancers 14, 4662 (2022).

48. Xie, Y.-H. et al. Fecal *Clostridium symbiosum* for Noninvasive Detection of Early and Advanced Colorectal Cancer: Test and Validation Studies. EBioMedicine 25, 32–40 (2017).

49. Liang, L. et al. Distinct microbes, metabolites, and the host genome define the multi-omics profiles in right-sided and left-sided colon cancer. Microbiome 12, 274 (2024).

50. Yang, Y. et al. Dysbiosis of human gut microbiome in young-onset colorectal cancer. Nat Commun 12, 6757 (2021).

51. Miquel, S., et al. *Faecalibacterium prausnitzii* and human intestinal health. Current Opinion in Microbiology 16, 255–261 (2013).

52. Sokol, H. et al. Faecalibacterium prausnitzii is an anti-inflammatory commensal bacterium identified by gut microbiota analysis of Crohn disease patients. Proceedings of the National Academy of Sciences 105, 16731–16736 (2008).

53. Díaz-Gay, M. et al. Geographic and age variations in mutational processes in colorectal cancer. Nature 643, 230–240 (2025).

54. Lee-Six, H. et al. The landscape of somatic mutation in normal colorectal epithelial cells. Nature 574, 532–537 (2019).

55. de Klaver, W. et al. Polyketide synthase positive Escherichia coli one-time measurement in stool is not informative of colorectal cancer risk in a screening setting. The Journal of Pathology 263, 217–225 (2024).

56. Chenard, T., Malick, M., Dube, J. & Masse, E. The influence of blood on the human gut microbiome. BMC Microbiol 20, 44 (2020).

57. Flemer, B. et al. The oral microbiota in colorectal cancer is distinctive and predictive. Gut 67, 1454–1463 (2018).

58. Kwong, T. N. Y. et al. Association Between Bacteremia From Specific Microbes and Subsequent Diagnosis of Colorectal Cancer. Gastroenterology 155, 383–390 e8 (2018).

59. Dai, Z. et al. Multi-cohort analysis of colorectal cancer metagenome identified altered bacteria across populations and universal bacterial markers. Microbiome 6, 70 (2018).

60. Lawrence, G. W. et al. A gut-derived Streptococcus salivarius produces the novel nisin variant designated nisin G and inhibits Fusobacterium nucleatum in a model of the human distal colon microbiome. mBio 16, e01573–24 (2024).

61. Li, Q. et al. Streptococcus thermophilus Inhibits Colorectal Tumorigenesis Through Secreting β-Galactosidase. Gastroenterology 160, 1179–1193.e14 (2021).

62. Jackson, M. A. et al. Proton pump inhibitors alter the composition of the gut microbiota. Gut 65, 749–756 (2016).

63. Nagata, N. et al. Population-level Metagenomics Uncovers Distinct Effects of Multiple Medications on the Human Gut Microbiome. Gastroenterology 163, 1038–1052 (2022).

64. Gacesa, R. et al. Environmental factors shaping the gut microbiome in a Dutch population. Nature 604, 732–739 (2022).

65. Istvan, P. et al. Exploring the gut DNA virome in fecal immunochemical test stool samples reveals associations with lifestyle in a large population-based study. Nat Commun 15, 1791 (2024).

66. Blanco-Míguez, A. et al. Extending and improving metagenomic taxonomic profiling with uncharacterized species using MetaPhlAn 4. Nat Biotechnol 41, 1633–1644 (2023).

67. Beghini, F. et al. Integrating taxonomic, functional, and strain-level profiling of diverse microbial communities with bioBakery 3. Elife 10, (2021).

68. Kieser, S., Brown, J., Zdobnov, E. M., Trajkovski, M. & McCue, L. A. ATLAS: a Snakemake workflow for assembly, annotation, and genomic binning of metagenome sequence data. BMC Bioinformatics 21, 257 (2020).

69. Bushnell, B. BBMap: BBMap short read aligner, and other bioinformatic tools. SourceForge https://sourceforge.net/projects/bbmap/ (2022).

70. Nurk, S., Meleshko, D., Korobeynikov, A. & Pevzner, P. A. metaSPAdes: a new versatile metagenomic assembler. Genome Res 27, 824–834 (2017).

71. Sieber, C. M. K. et al. Recovery of genomes from metagenomes via a dereplication, aggregation and scoring strategy. Nat Microbiol 3, 836–843 (2018).

72. Kang, D. D. et al. MetaBAT 2: an adaptive binning algorithm for robust and efficient genome reconstruction from metagenome assemblies. PeerJ 7, e7359 (2019).

73. Wu, Y.-W., Simmons, B. A. & Singer, S. W. MaxBin 2.0: an automated binning algorithm to recover genomes from multiple metagenomic datasets. Bioinformatics 32, 605–607 (2016).

74. Olm, M. R., Brown, C. T., Brooks, B. & Banfield, J. F. dRep: a tool for fast and accurate genomic comparisons that enables improved genome recovery from metagenomes through de-replication. ISME J 11, 2864–2868 (2017).

75. Chaumeil, P.-A., Mussig, A. J., Hugenholtz, P. & Parks, D. H. GTDB-Tk: a toolkit to classify genomes with the Genome Taxonomy Database. Bioinformatics 36, 1925–1927 (2020).

76. Willett, W., Howe, G. & Kushi, L. Adjustment for total energy intake in epidemiologic studies. The American Journal of Clinical Nutrition 65, 1220S–1228S (1997).

77. Barrubés, L. et al. Association between the 2018 WCRF/AICR and the Low-Risk Lifestyle Scores with Colorectal Cancer Risk in the Predimed Study. Journal of Clinical Medicine 9, 1215 (2020).

78. Onyeaghala, G. et al. Adherence to the World Cancer Research Fund/American Institute for Cancer Research cancer prevention guidelines and colorectal cancer incidence among African Americans and whites: The Atherosclerosis Risk in Communities study. Cancer 126, 1041–1050 (2020).

79. Petimar, J. et al. Adherence to the World Cancer Research Fund/American Institute for Cancer Research 2018 Recommendations for Cancer Prevention and Risk of Colorectal Cancer. Cancer Epidemiology, Biomarkers & Prevention 28, 1469–1479 (2019).

80. Wang, P., Song, M., Eliassen, A. H., Wang, M. & Giovannucci, E. L. Dietary patterns and risk of colorectal cancer: a comparative analysis. International Journal of Epidemiology 52, 96–106 (2023).

81. WHO Collaborating Centre for Drug Statistics Methodology. ATC index with DDDs 2022. WHO Collaborating Centre, Oslo.

82. World Cancer Research Fund International & American Institute for Cancer Research. Dietary and Lifestyle Patterns for Cancer Prevention: Evidence and Recommendations from CUP Global. https://www.wcrf.org/wp-content/uploads/2025/04/DLP_Full_Report_FINAL.pdf (2025).

83. vegan: Community Ecology Package. R package version 2.5-7. (2020).

84. Kuhn, M. & Wickham, H. Tidymodels: a collection of packages for modeling and machine learning using tidyverse principles. (2020).

85. He, H., Bai, Y., Garcia, E. A. & Li, S. ADASYN: Adaptive synthetic sampling approach for imbalanced learning. in 2008 IEEE International Joint Conference on Neural Networks (IEEE World Congress on Computational Intelligence) 1322–1328 (2008). doi:10.1109/IJCNN.2008.4633969.

86. Mallick, H. et al. Multivariable association discovery in population-scale meta-omics studies. PLOS Computational Biology 17, e1009442 (2021).

