## Supplementary information for "Microbiome signatures for detection of colorectal lesions in population-based FIT screening"

### Supplementary data

**Supplementary Data 1:** Host variable associations with gut microbiome alpha-diversity.

**Supplementary Data 2:** Host variable associations with gut microbiome beta diversity based on Bray-Curtis distances.

**Supplementary Data 3:** Overview of species associations with outcome groups.

**Supplementary Data 4:** Overview of KO associations with outcome groups.

**Supplementary Data 5:** Species associations with participant characteristics. Species included are those that were significantly associated with outcome measures.

**Supplementary Data 6:** Overview of prevalence for selected species that have previously been associated with CRC in other studies.

**Supplementary Data 7:** List of the diet, lifestyle, and demography variables selected for inclusion.

### Supplementary tables

**Supplementary Table 1**: Overview of clinicopathological diagnoses of individuals whose samples were not included in sequencing.

**Supplementary Table 2**: Participant characteristics by outcome categories.

**Supplementary Table 3**:
Overview of drug use prevalence in: CRCbiome prior to inclusion in BCSN; the remaining FIT arm of BCSN study participants; in a nationally representative screening-eligible population; and in the CRCbiome population prior to sample collection for CRCbiome.

**Supplementary Table 4**: Odds ratios (ORs) and 95% confidence intervals (CIs) for outcome group with a given microbial community state, compared with all other community states.

**Supplementary Table 5**: Overview of all lesion localizations from endoscopist records.

**Supplementary Table 6**: Significantly differential gene carriage in *Faecalibacterium* bacteria according to the presence of distal lesions.

### Supplementary figures

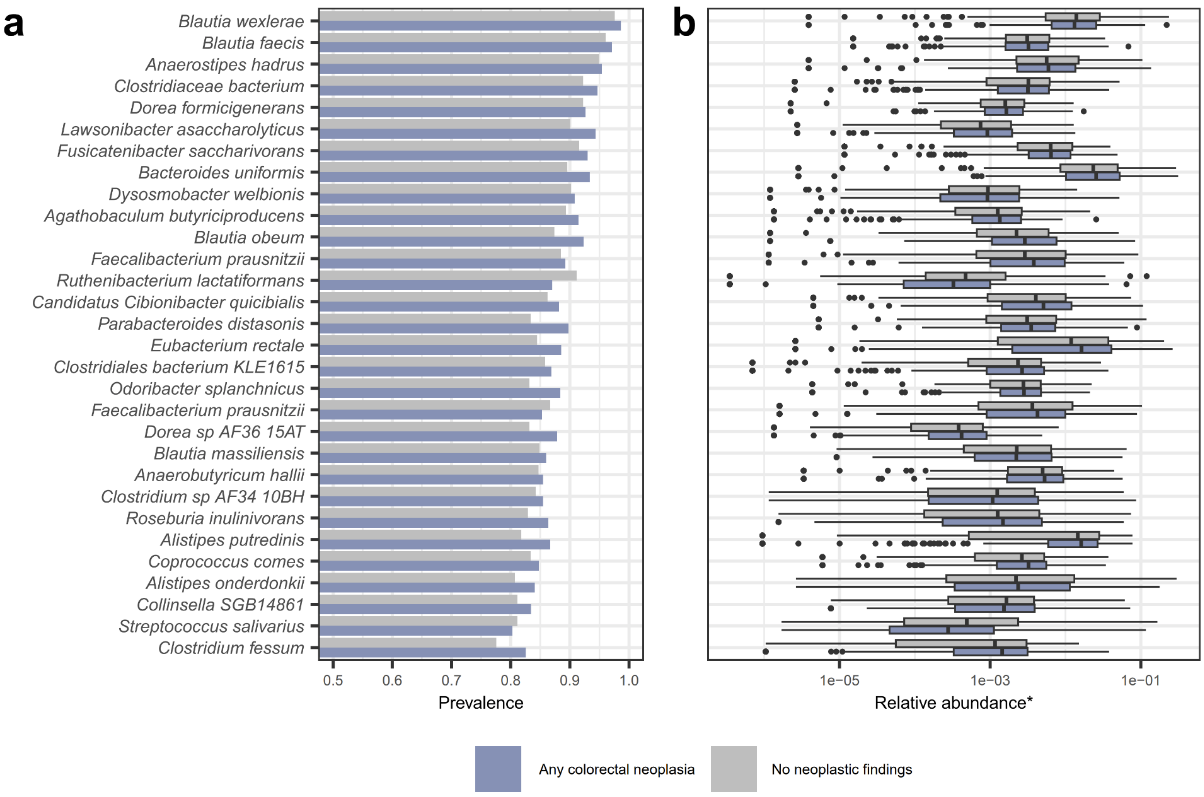


**Supplementary Figure S1**: Common species in the CRCbiome dataset. Prevalence (a) and relative abundance (b) of the 30 most prevalent microbial species stratified by outcome category defined any colorectal neoplasia (blue) or no neoplastic findings (gray). For visualization purposes, relative abundance of zero has been omitted.

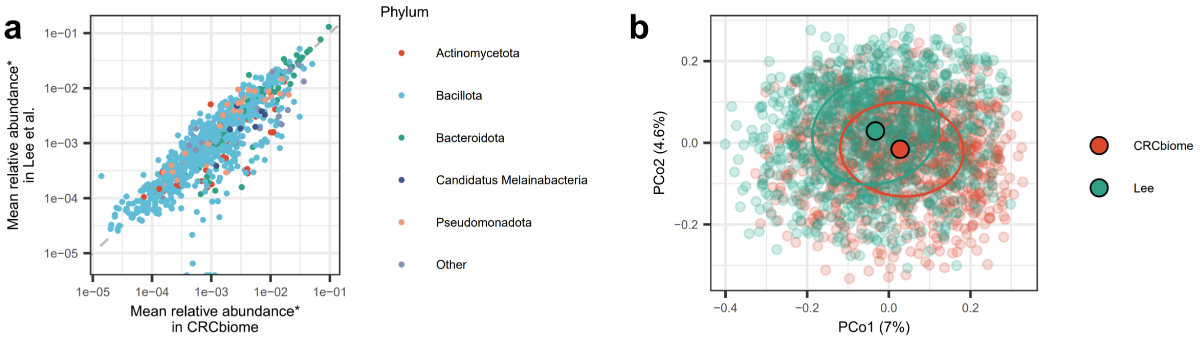


**Supplementary Figure S2**: Microbial composition of CRCbiome and Lee et al.[1] a) Mean relative abundance for CRCbiome participants and in the Lee et al. dataset, for any species found in more than 5% of samples in either dataset. Three species with > 5% prevalence in CRCbiome were not identified in Lee et al. (points along x-axis). The dashed line indicates equal mean relative abundance. *abundance only considered when a species was present. b) PCoA based on Bray-Curtis distances of relative species abundance in CRCbiome and Lee et al. Ellipses encompass 50% of observations and solid points indicate group centroids.

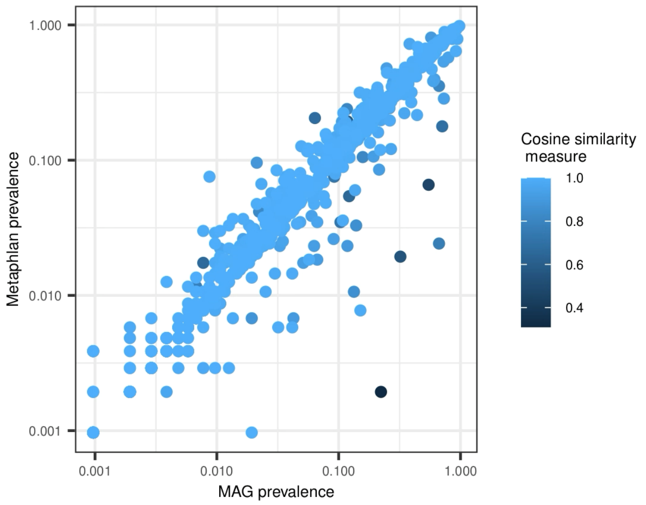


**Supplementary Figure S3**: Comparison of microbial profiles captured by MetaPhlAn and quantification of metagenome-assembled genomes (MAGs). Species annotated with identical names (using GTDB annotations) by the two profiling approaches are included in the comparison. This included 819 species, out of 2609 annotated species detected by MetaPhlAn and 1208 unique species annotated for the MAGs. The Spearman correlation coefficient for species prevalence measurements between the two profiling methods was 0.97, and 92.7% of species had a cosine similarity measure over 0.95.

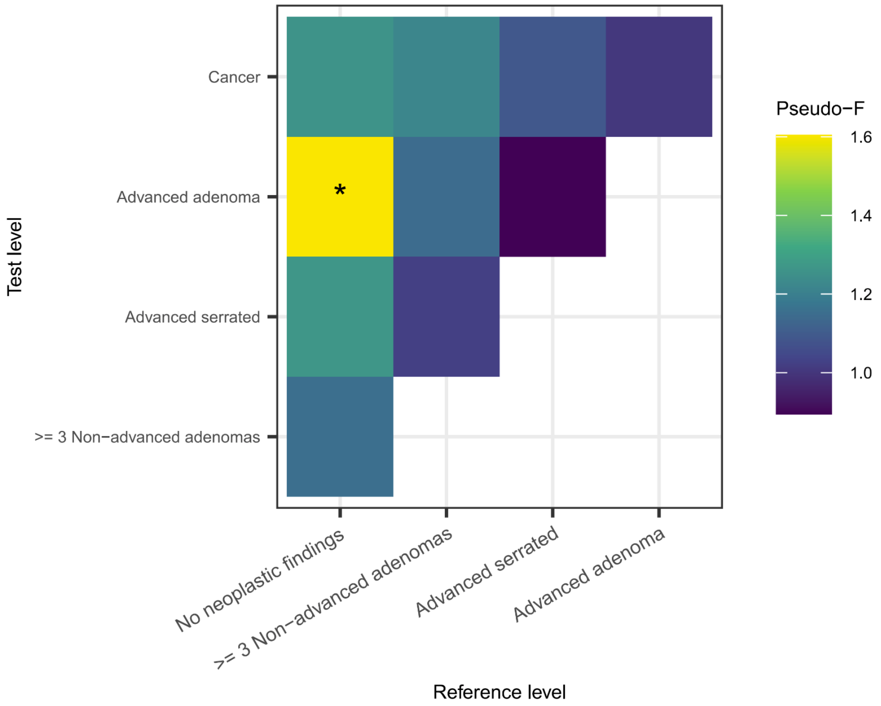


**Supplementary Figure S4**: Pairwise comparison of within- and between-group variance in gut microbiome beta diversity for participants based on outcome category. The ratio of within-group variance to between-group variance was measured using the pseudo-F statistic, with significance (unadjusted p-value < 0.05) in pairwise testing indicated by an asterisk. Statistical tests were performed using PERMANOVA as implemented in the adonis2 function, adjusted for sex, age, screening center, sequencing depth, the WCRF score, smoking status, level of education, findings of hemorrhoids, findings of IBD, and recent antibiotics.

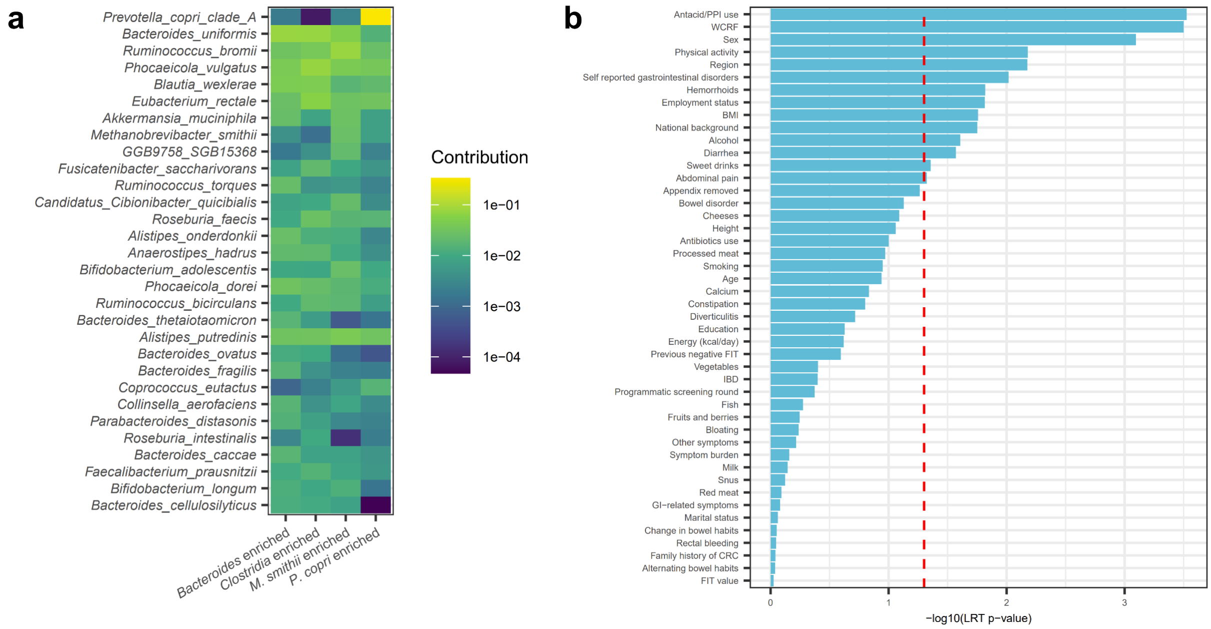

**Supplementary Figure S5**: Dirichlet multinomial mixture models[2] (DMMs) based community state classification of CRCbiome participants. a) The contribution of species to each component of the DMM for the 30 species with the highest absolute deviations across components from a single-component model. b) Significance of multinomial logistic regression models assessing associations between metagenome community state and host factors (y-axis), adjusting for sex, age, and screening center.

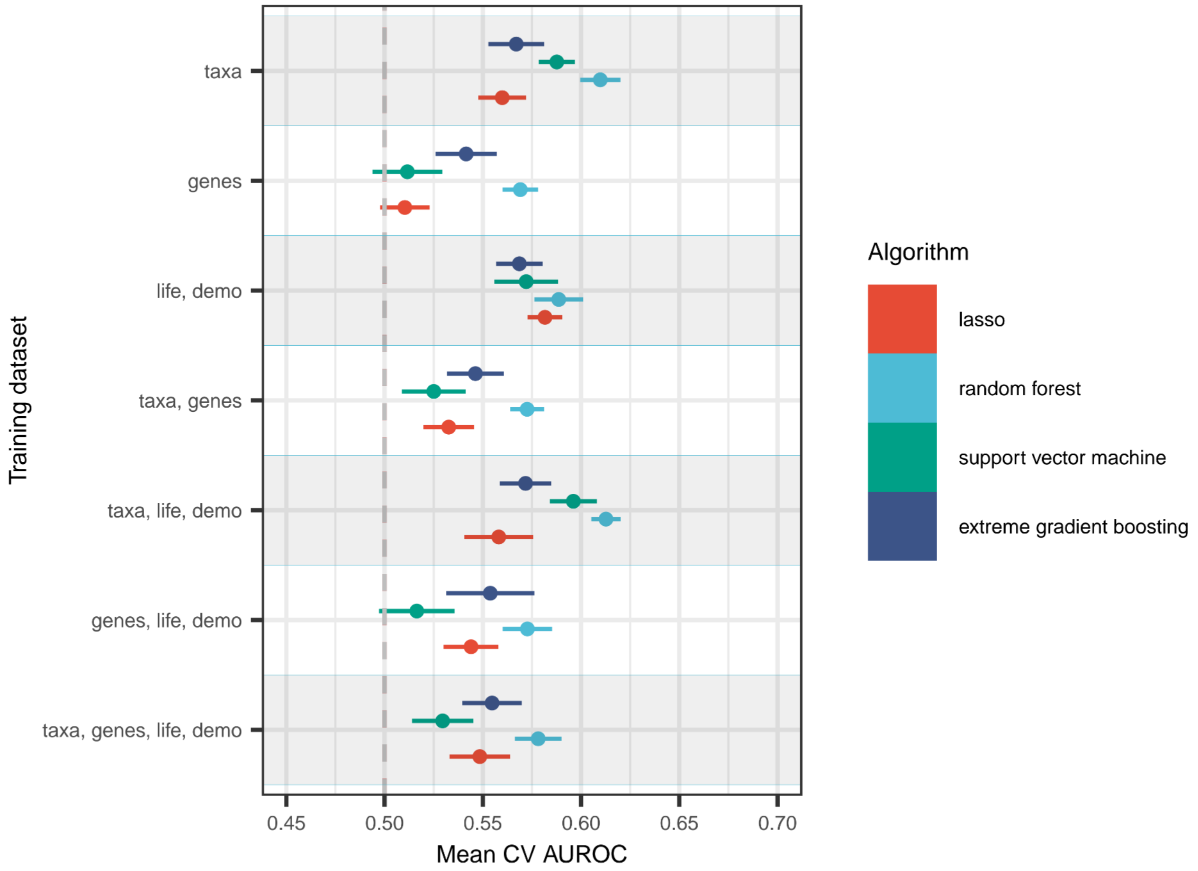


**Supplementary Figure S6**: Comparison of machine learning algorithms. The performance of four machine learning algorithms in distinguishing screening participants with or without CRC-related findings at colonoscopy was assessed using 20 times repeated 5-fold cross validation. Input datasets included microbial species abundances (taxa), microbial gene defined by KEGG orthogroups (genes), or a set of demographic and lifestyle factors (see Supplementary Table 6).

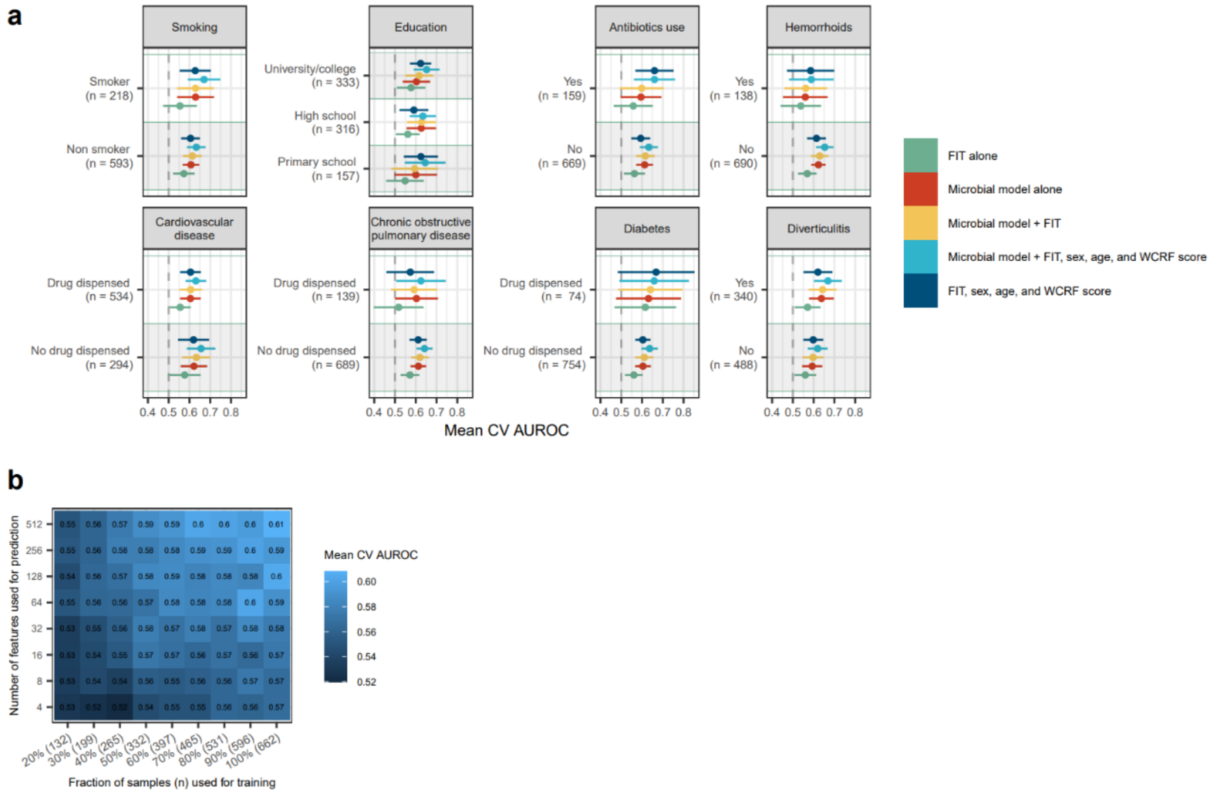


**Supplementary Figure S7**: Classification of any colorectal neoplasia by participant characteristics, and the effect of feature selection and sample size. a) Predictive performance where predictions are stratified by participant characteristics. Drug use during the year leading up to screening sample collection as an indication of comorbidities (as reported in supplementary table S2) were used for stratification in the right-hand panels. b) Random forest models were set up for prediction of any colorectal neoplasia, following pre-selection of a defined number of features (y-axis) based on importance measures from random forest models, using a defined fraction of the training set as input to the cross-validation procedure (5 times repeated, 5-fold cross validation).

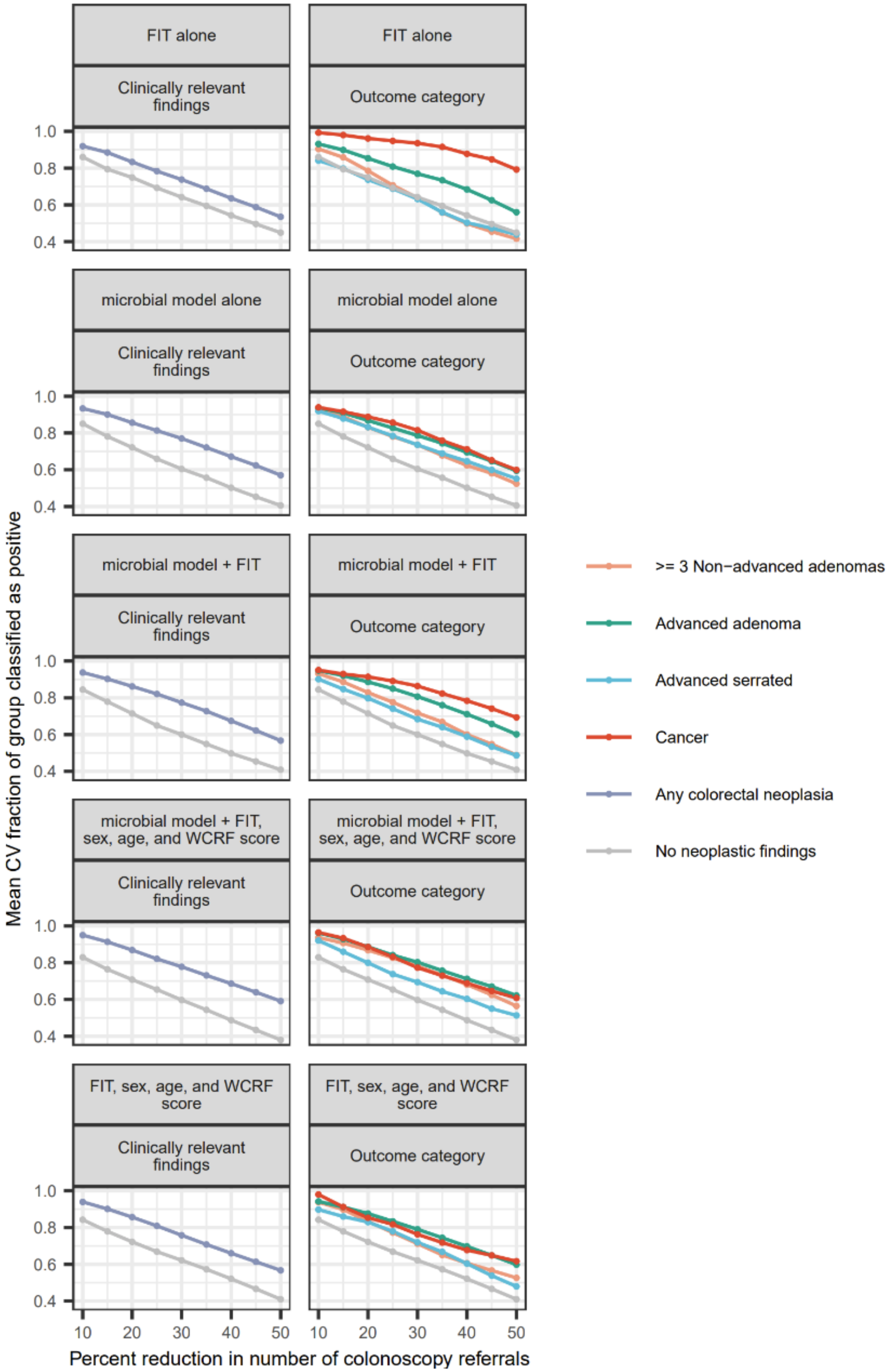

FIT alone
Clinically relevant findings
Outcome category
microbial model alone
microbial model + FIT
microbial model + FIT, sex, age, and WCRF score
FIT, sex, age, and WCRF score
1.0
0.8
0.6
0.4
10
20
30
40
50
Percent reduction in number of colonoscopy referrals
Mean CV fraction of group classified as positive
>= 3 Non−advanced adenomas
Advanced adenoma
Advanced serrated
Cancer
Any colorectal neoplasia
No neoplastic findings

**Supplementary Figure S8**: ML-based classification by positivity threshold. Thresholds were defined by positivity rates in validation sets to assign a certain fraction of cases to be "positive". The fraction of cases thus classified as negative was used as a projection of a potential reduction in colonoscopy referrals, and the fraction of each outcome category classified as positive were enumerated for each threshold. Points indicate the mean of classification results across 20 times repeated 5-fold cross-validation.

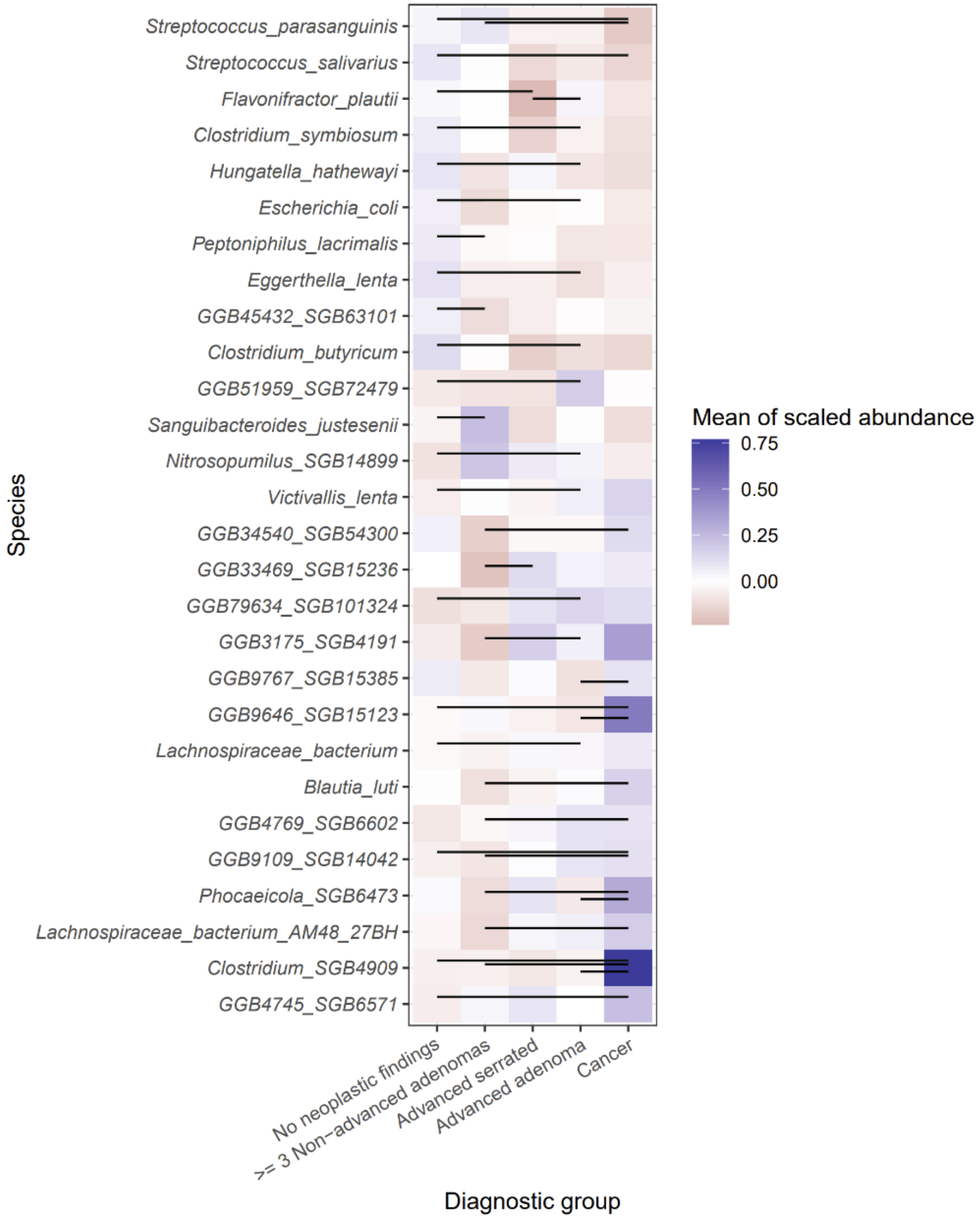

**Supplementary Figure S9**: Pairwise comparison of species abundance between diagnostic groups. For each species, the relative abundance was scaled to mean 0 and standard deviation of 1, with the mean for each group indicated by the color of each cell. Significant pairwise differences in abundance (FDR < 0.05) are indicated by lines. Species with significant difference in any pairwise comparison are shown (n = 28), arranged according to the effect size of difference between no neoplastic findings and cancer.

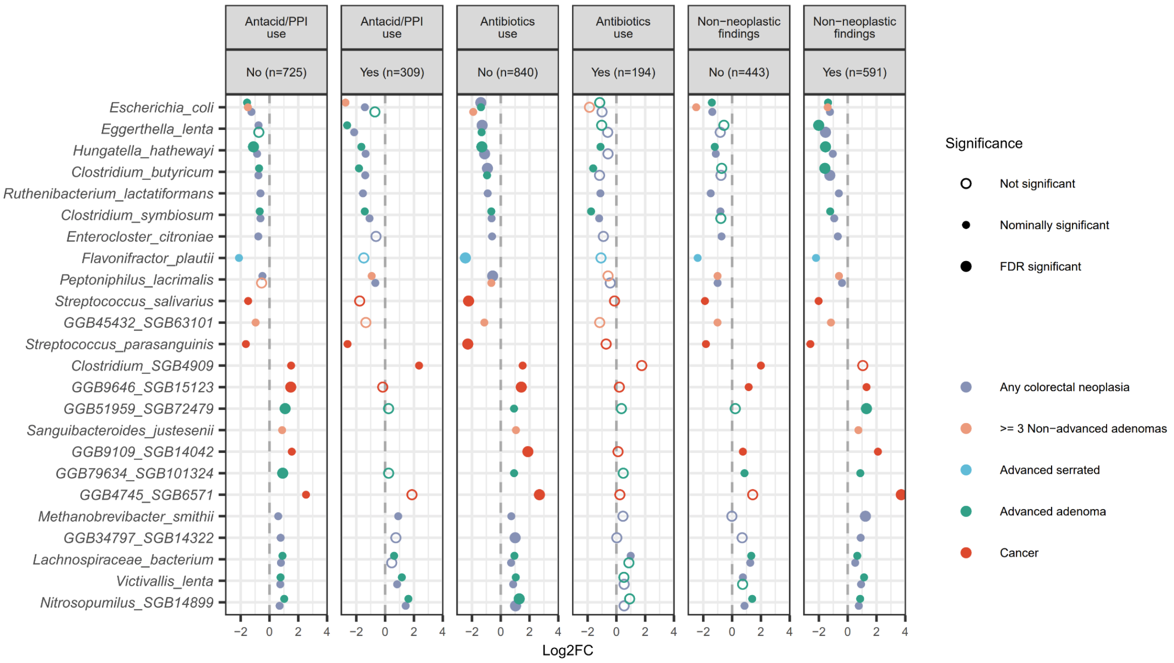


**Supplementary Figure S10**: Sensitivity analyses for differential abundance of microbial species. Separate differential abundance models were set up for subgroups of participants; results are reported in one panel per subgroup. Participants with non-neoplastic findings had findings of diverticulitis, hemorrhoids, IBD, or other conditions not related to CRC uncovered at colonoscopy. Models were adjusted as described for Figure 4a), except for those without non-neoplastic, which were not adjusted for the presence of hemorrhoids or IBD.

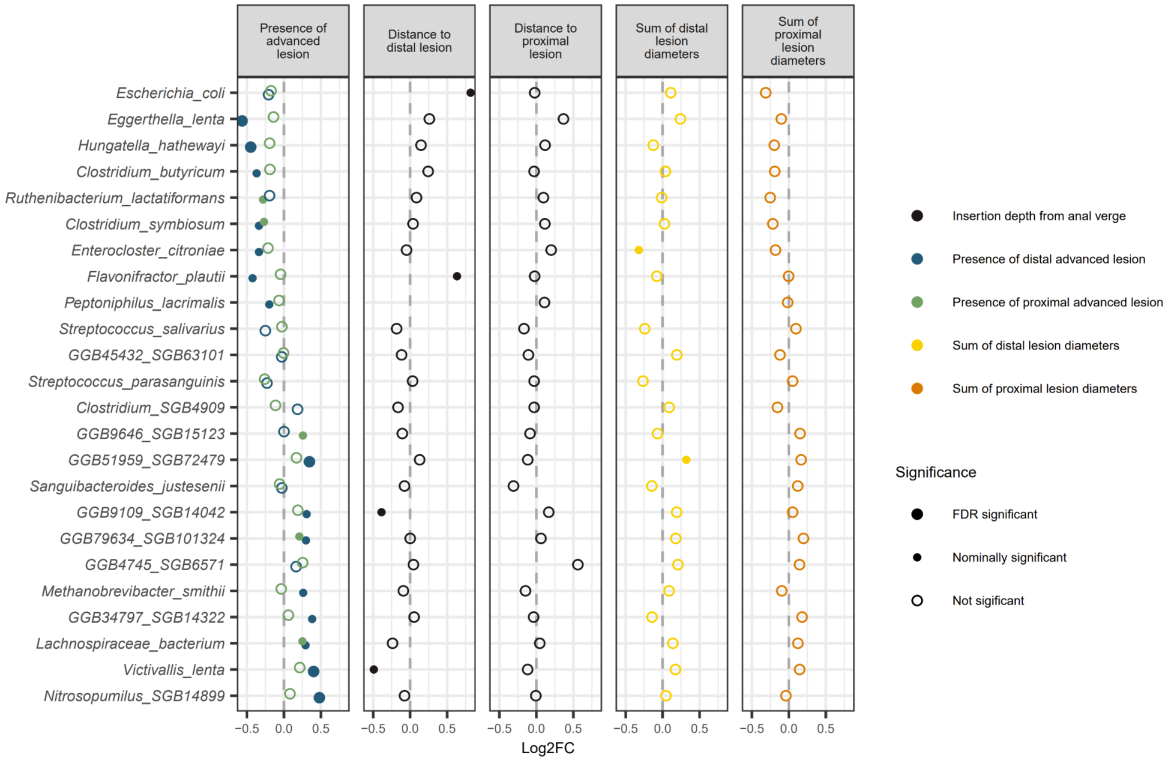


**Supplementary Figure S11**: Differentially abundant species according to lesion localization and lesion load. Species with differential abundance according to outcome category (as presented in Figure 4) were assessed for differential abundance in relation to lesion localization and lesion load using five alternative models. In panel one from the left, the presence of advanced colorectal lesions in the proximal and distal portions of the large intestine were included as independent variables. In panel three and four, the distance to lesion considered most serious were included in models where only individuals with any colorectal neoplasia in the distal or proximal regions were included, respectively. In panel five and six, the load of lesions was included as independent variables, with one model for those with a distal lesion load of more than zero, and one for those with a proximal lesion load of more than zero.
